# Health behaviours the month prior to COVID-19 infection and the development of self-reported long COVID and specific long COVID symptoms: A longitudinal analysis of 1,811 UK adults

**DOI:** 10.1101/2022.04.12.22273792

**Authors:** Elise Paul, Daisy Fancourt

## Abstract

**Background:** Demographic and infection-related characteristics have been identified as risk factors for long COVID, but research on the influence of health behaviours (e.g., exercise, smoking) immediately preceding the index infection is lacking.

**Methods:** 1,811 UK adults from the UCL COVID-19 Social Study and who had previously been infected with COVID-19 were analysed. Health behaviours in the month before infection were weekly exercise frequency, days of fresh air per week, sleep quality, smoking, consuming more than the number of recommended alcoholic drinks per week (>14), and the number of mental health care behaviours (e.g., online mental health programme). Logistic regressions controlling for covariates (e.g., COVID-19 infection severity and pre-existing health conditions) examined the impact of health behaviours on long COVID and three long COVID symptoms (difficulty with mobility, cognition, and self-care).

**Results:** In the month before infection with COVID-19, poor quality sleep increased the odds of long COVID (odds ratio [OR]: 3.53; (95% confidence interval [CI]: 2.01 to 6.21), as did average quality sleep (OR: 2.44; 95% CI: 1.44 to 4.12). Having smoked (OR: 8.39; 95% CI: 1.86 to 37.91) increased and meeting recommended weekly physical activity guidelines (3+ hours) (OR: 0.05; 95% CI: 0.01 to 0.39) reduced the likelihood of difficulty with self-care (e.g., washing all over or dressing) amongst those with long COVID.

**Conclusion:** Results point to the importance of sleep quality for long COVID, potentially helping to explain previously demonstrated links between stress and long COVID. Results also suggest that exercise and smoking may be modifiable risk factors for preventing the development of difficulty with self-care.

**Funding:** The Nuffield Foundation [WEL/FR-000022583], the MARCH Mental Health Network funded by the Cross-Disciplinary Mental Health Network Plus initiative supported by UK Research and Innovation [ES/S002588/1], and the Wellcome Trust [221400/Z/20/Z and 205407/Z/16/Z].

**What is already known on the topic:** Long COVID is rapidly becoming a public health concern. Although existing evidence to date has identified health characteristics such as obesity as risk factors, hardly any research on modifiable risk factors such as health behaviours has been conducted.

**What this study adds:** This study adds to the dearth of evidence on modifiable risk factors occurring before COVID-19 infection. Findings suggest a role of poor sleep quality for the development of long COVID, and for meeting physical activity guidelines (3+ hours per week) and not smoking as modifiable risk factors for self-care difficulties amongst those with long COVID.

## Introduction

Long COVID, which includes both ongoing symptomatic COVID-19 (the presence of symptoms from 4 to 12 weeks post-onset), and post-COVID-19 syndrome (the presence of symptoms > 12 weeks post-onset)^1^ is rapidly becoming a major public health concern.^2^ Findings from studies representative of the general adult population in England suggest that around one in five (19%- 22%) of people previously infected with COVID-19 self-report long COVID,^3,4^ with over half (67%) reporting that their ongoing symptoms significantly impacted their ability to carry out their day to day activities.^3^ The most common symptoms are weakness, fatigue, cognitive difficulties (e.g., concentration and remembering), and breathlessness.^3,5,6^ Additionally, a large portion of patients suffering from long COVID report reduced quality of life.^5,6^

There is some emerging evidence of the prominent role of pre-infection health factors in the development and experience of long COVID. For example, being obese or overweight is associated with risk of long COVID,^4,7–10^ likely via mechanisms including chronic systemic inflammation.^11^ However, work on behavioural risk factors for long COVID remains in its infancy. Several lifestyle factors, including smoking^12^ physical inactivity,^13^ poor sleep^14,15^ and excessive alcohol consumption not only increase the risk of infectious diseases, but can also impede vaccine response.^11^ One study found smoking to increase the likelihood of persistent COVID symptoms (12 weeks or more).^4^ However, findings were cross-sectional and analyses did not include other health behaviours which may influence long COVID. It therefore remains unclear whether other behavioural factors influence risk of developing long COVID.

This is important to ascertain, as negative changes have been observed in many of these health behaviours during the pandemic. For example, in England, the prevalence of higher risk alcohol consumption has increased, and physical activity levels have declined,^16^ whilst sleep difficulties have also increased.^17^ If such behavioural factors are found to be risk factors for long COVID it could help inform public health programmes designed to reduce the risk of further cases of long COVID both during this pandemic and in potential future pandemics. Thus, the aim of this study was to identify whether specific health behaviours present in the month preceding infection with COVID-19 act as upstream and potentially modifiable markers of long COVID.

## Methods

### Study design and participants

Data were drawn from the COVID-19 Social Study; a large panel study of the psychological and social experiences of over 75,000 adults (aged 18+) in the UK during the COVID-19 pandemic. The study commenced on 21 March 2020 and involves online data collection from participants for the duration of the COVID-19 pandemic in the UK. Data were initially collected weekly (from study commencement through August 2020), then monthly thereafter. The study is not random and therefore is not representative of the UK population. But it does contain a well-stratified sample that was recruited using three primary approaches. First, convenience sampling was used, including promoting the study through existing networks and mailing lists (including large databases of adults who had previously consented to be involved in health research across the UK), print and digital media coverage, and social media. Second, more targeted recruitment was undertaken focusing on groups who were anticipated to be less likely to take part in the research via our first strategy, including (i) individuals from a low-income background, (ii) individuals with no or few educational qualifications, and (iii) individuals who were unemployed. Third, the study was promoted via partnerships with third sector organisations to vulnerable groups, including adults with pre-existing mental health conditions, older adults, carers, and people experiencing domestic violence or abuse. The study was approved by the UCL Research Ethics Committee [12467/005] and all participants gave informed consent. Participants were not compensated for participation (https://osf.io/jm8ra/). We included participants who met the five criteria outlined in Figure 1. First, participants were included if they had participated in the November 2021 survey and said that they had at some prior point been infected with COVID-19. Second, the date given for their COVID-19 infection had to be non-missing and had to be no earlier than 27 April 2020 and at least 5 weeks prior to completion of the specific questions on long COVID. 27 April 2020 was chosen as we were interested in health behaviours in the month prior to COVID-19 infection, and the collection of all individual items comprising these variables commenced 13 April 2020. Five weeks was chosen as the minimum time period as many studies on long COVID apply a threshold of “more than four weeks of symptoms” to be experienced for the term long COVID to be applied.^8,18,19^ Third, participants who had had COVID-19 only once were included; participants who reported more than one infection were excluded to avoid overlapping symptoms from the two infections. Fourth participants had to have participated in the study in the month prior to the date of their infection to gather health behaviour data. Fifth, participants had to have non-missing data on long COVID outcome variables (presence/absence and specific long COVID symptoms) and study variables required to calculate statistical weights (gender, age, ethnicity, country, and education). The final analytic sample comprised 1,811. See Figure 1 for a flow chart of the number of participants excluded at each step.

**Figure 1.**
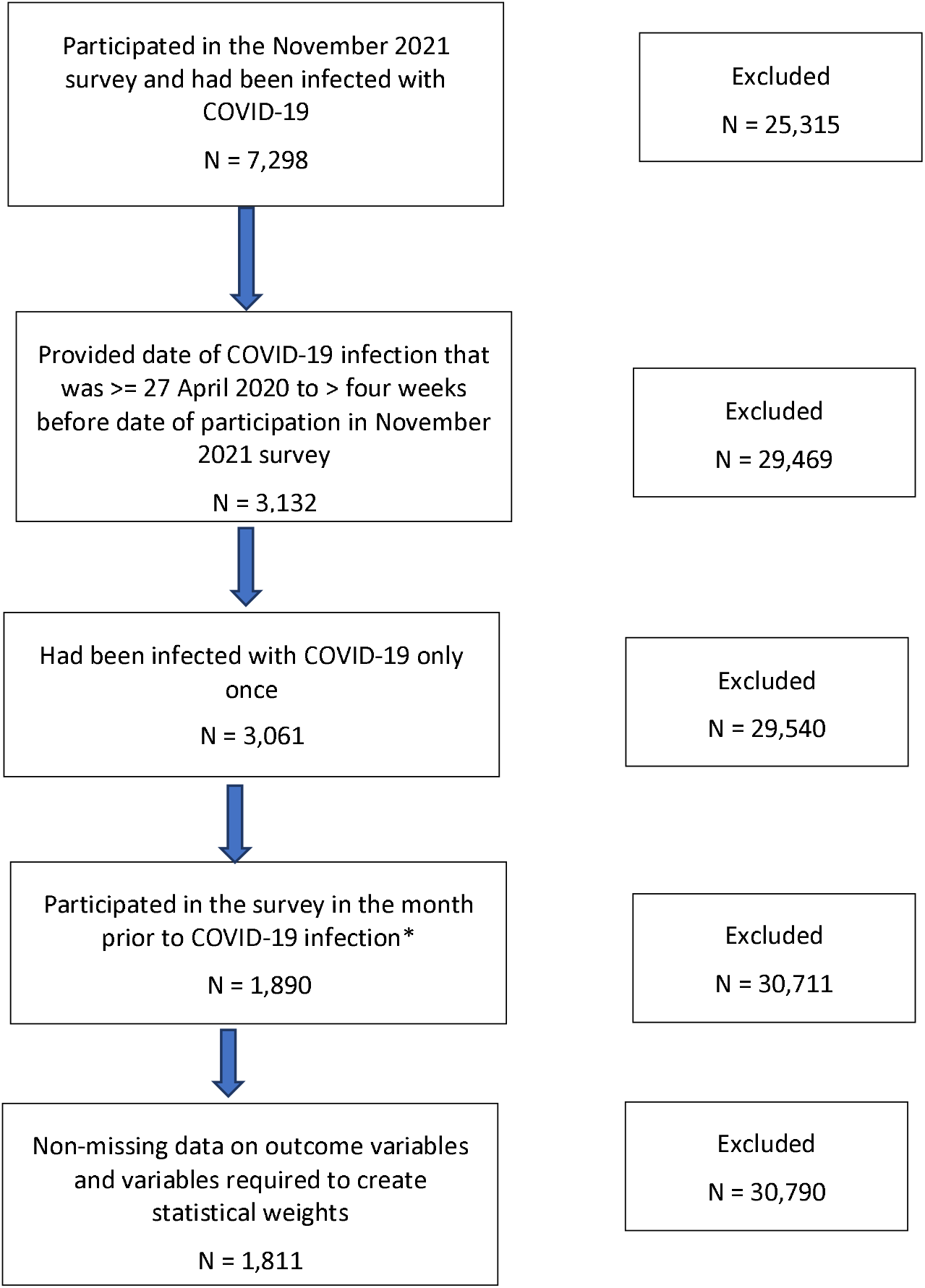
Flow chart of sample selection. *Note. Data from two weeks prior to COVID-19 infection were used, and when unavailable, consecutively further weeks prior, up to six weeks.

We used multiple imputation by chained equations to generate 50 imputed datasets for participants who met all study inclusion criteria but had missing data on other study variables (Supplemental Table S1). Imputation models included all variables used in the analysis, as well as additional auxiliary variables (e.g., home ownership status, depressive symptoms at baseline). Substantive results using cases without any missing data and the imputed sample were similar (Supplemental Tables S2–S5). See Supplemental Table S6 for a comparison of excluded and included participants on study variables.

## Measures

### Outcome variables

The presence of long COVID was measured with response to the question: “Do you consider yourself to have (or have had) Long Covid?”. The four response options (Supplemental Table S7) were categorised into (i) no and (ii) yes (formally diagnosed or suspected). Sensitivity analyses were carried out to test whether results were consistent when including participants who were “unsure” about whether they had had long COVID within the case group.

To look at the presence of three specific long COVID symptoms, we restricted the sample to people with long COVID. These three variables were operationalised from questions assessing the extent to which participants had difficulty with (i) mobility, (ii) cognition, and (iii) self-care (Supplemental Table S7). Response options were on a four-point scale ranging from 0 “no difficulty at all” to “unable to do” and treated as binary (present vs absent) in analyses due to low numbers within response categories.

### Predictor variables

#### Health behaviours

Six health behaviour variables constructed from study-developed questions were considered (Supplemental Table S7). Data starting with two weeks before the COVID-19 infection were used, and if unavailable, data from three weeks, then four, up to six weeks. See Supplemental Table S8 for the distribution of data weeks used. Frequency of exercise undertaken in a single week was operationalised as none vs <30 minutes to two hours vs three hours or more. We selected these categories as the latter reflects current weekly physical activity recommendations in the UK.^20^ For days outside, a count of the number of days participants had left the house in the past week for at least 15 minutes was included. Sleep quality in the past week was operationalised as very good/good vs average vs not good/very poor. Smoking (non-smoker vs any smoking), and a binary variable indicating 14 or more alcoholic drinks vs fewer than 14 alcoholic drinks in the past week were also included. Fourteen was chosen as the cut-off for alcohol consumption to reflect current recommendations on alcohol intake per week in the UK.^21^ Finally, the number of mental health care behaviours was included (e.g., taken medications, spoke to somebody on a support line; Supplemental Tables S7). Because increasing weight and obesity are associated with long COVID,^7–9^ and are also risk factors for chronic disease independent of physical activity,^22^ we conducted sensitivity analyses with a variable reflecting overweight status collected in June 2020 (slightly underweight or normal weight vs slightly overweight or very overweight).

### Covariates

#### COVID-19 infection variables

COVID-19 infection severity was assessed with the study developed question: “How severe were your symptoms in the first 1-2 weeks?”. Response options were categorised into (i) asymptomatic, (ii) mild (experienced symptoms but was able to carry on with daily activities), (iii) moderate (experienced symptoms and had to rest in bed), and (iv) severe (participant was hospitalised).

A variable to indicate which strain of the virus was dominant in the UK^23^ at the time of infection was included and coded as (0) the original COVID-19 variant (31 January to 31 October 2020, (1) Alpha (1 November 2020 to 30 June 2021), (2) Delta (1 July 2021 to 30 November 2021), and (3) Omicron (1 December 2021 onwards).

#### Socio-demographics

Socio-demographic factors were collected at baseline and included gender (male vs female), age group (60+, 45-59, 30-44, and 18-29) ethnicity (white vs ethnic minority groups [i.e., Asian/Asian British, Black/Black British etc. See Supplemental Table S7 for a full listing of response options]), education (undergraduate degree or higher, A-levels (equivalent to education to age 18) or vocational training, and up to GCSE (General Certificate of Secondary Education) [equivalent to education to age 16]), low income (annual household income: <£30,0000), employment status (not employed [still at school, at university, unable to work due to disability, unemployed and seeking work, or retired vs employed [self-employed, part-time or full-time employed]), government’s identified key worker status (vs not a key worker), crowded household (< one room per person), living arrangement (living alone vs living with others but not including children vs living with others, including children), and area of dwelling (urban [city, large town, small town] vs rural [village, hamlet, isolated dwelling]). Key workers included people with jobs deemed essential during the pandemic (e.g., health and social care, education) and who were required to leave home to carry out this work during lockdowns.

#### Pre-existing health conditions

Participants reported whether they had received a clinical diagnosis of a mental health condition (e.g., depression, anxiety, or another clinically diagnosed mental health problem) or chronic physical health condition (e.g., high blood pressure, diabetes, lung disease (e.g., asthma or COPD)). Responses were used to create two binary variables to indicate the presence of pre-existing physical and mental health conditions.

### Statistical analysis

Two sets of analyses were carried out. First, binary logistic regression models were fitted to examine associations of health behaviours in the month before infection with COVID-19 and the development of long COVID. Second, binary logistic regression models were fitted to examine associations between health behaviours in the month prior to COVID-19 infection and the presence of each of the three specific long COVID symptoms (difficulty with mobility, cognition, and self-care) amongst participants with long COVID.

For both sets of analyses, Model 1 included only health behaviours in the same model, Model 2 additionally adjusted for COVID-19 infection variables, Model 3 additionally adjusted for socio-demographic characteristics, and Model 4 additionally adjusted for pre-existing health conditions. Robust standard errors were used in all analyses. Coefficients from the binary logistic regressions were exponentiated and presented as odds ratios (OR), along with corresponding 95% confidence intervals (CI).

To account for the non-random nature of the sample and increase representativeness of the UK general population, all data were weighted to the proportions of gender, age, ethnicity, country, and education obtained from the Office for National Statistics.^24^ A multivariate reweighting method was implemented using the Stata user written command ‘ebalance’.^25^ Further details are shown in the User Guide (https://osf.io/jm8ra/). Analyses were conducted using Stata version 16.^26^

## Results

One in five (20.48%) either believed themselves to have or had been diagnosed with long COVID, and a further 12.04% were unsure (Supplemental Table S9). The most often reported long COVID symptom amongst those with long COVID was difficulty with cognition (62.58%), followed by difficulty with mobility (55.49 (Table 1). People living in crowded accommodation, who had a physical or mental health condition, who lived with children, had low levels of education or income, and had moderate or severe COVID-19 were all more likely to develop long COVID.

**Table 1.**
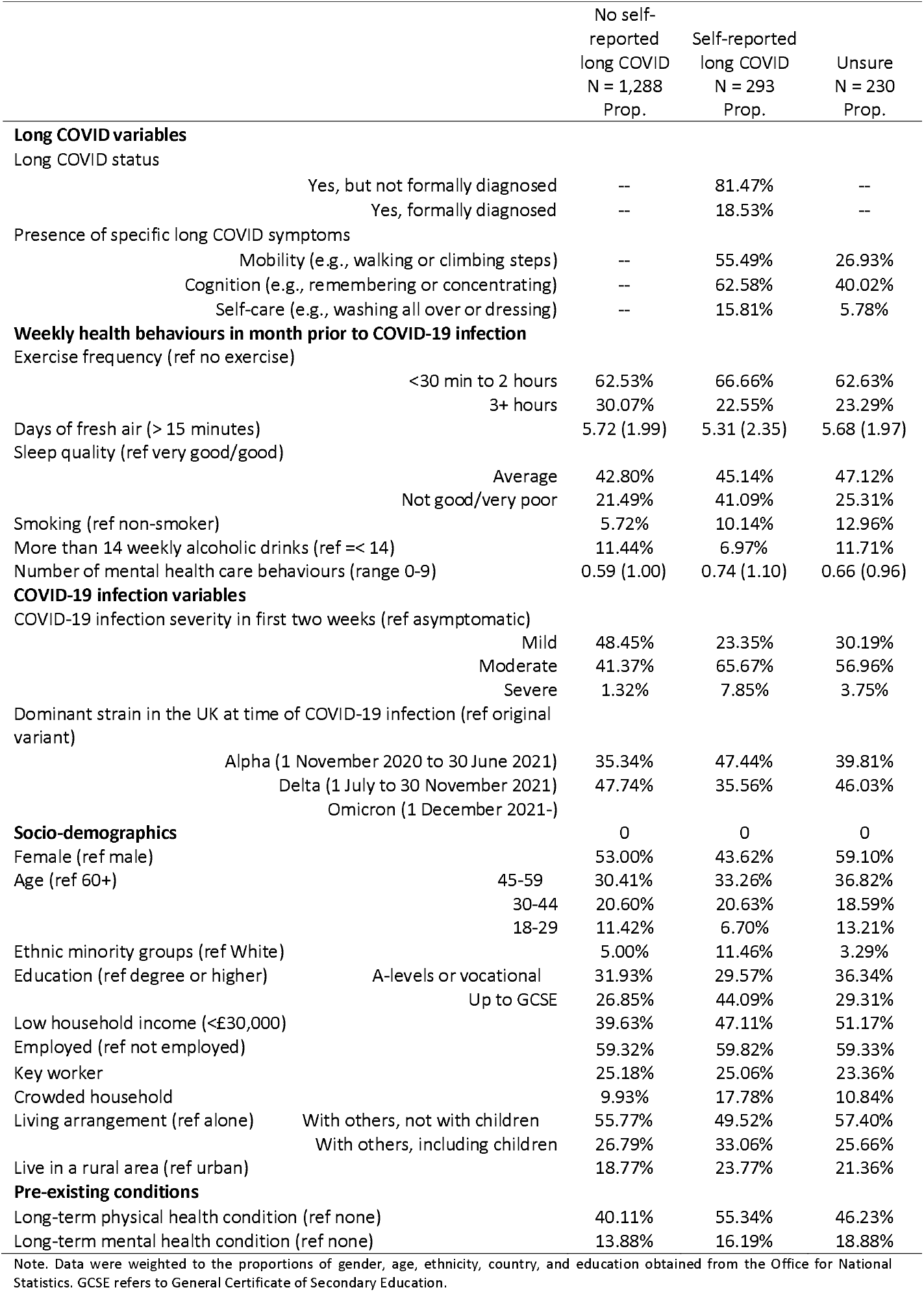
Descriptive characteristics of the sample by long COVID status, weighted (N =1,811)

**Table 2.** Descriptive characteristics of participants with long COVID by specific long COVID symptom, weighted (N = 293)

|  |  | Difficulty<br>with<br>mobility<br>N = 159<br>Prop. | Difficulty<br>with<br>cognition<br>N = 199<br>Prop. | Difficulty<br>with self-care<br>N = 45<br>Prop. |
| --- | --- | --- | --- | --- |
| **Long COVID variables** |  |  |  |  |
| Long COVID status |  |  |  |  |
|  | Yes, but not formally diagnosed | 73.42% | 76.61% | 62.14% |
|  | Yes, formally diagnosed | 26.58% | 23.39% | 37.86% |
| Presence of specific long COVID symptoms |  |  |  |  |
|  | Mobility (e.g., walking or climbing steps) | -- | 58.81% | 100.00% |
|  | Cognition (e.g., remembering or concentrating) | 66.33% | -- | 84.85% |
|  | Self-care (e.g., washing all over or dressing) | 28.49% | 21.44% | -- |
| **Weekly health behaviours in month prior to COVID-19 infection** |  |  |  |  |
| Exercise frequency (ref no exercise) |  |  |  |  |
|  | <30 min to 2 hours | 65.22% | 64.15% | 66.67% |
|  | 3+ hours | 20.29% | 24.83% | 10.37% |
| Days of fresh air (> 15 minutes) |  | 4.89 (2.54) | 5.19 (2.41) | 4.38 (2.31) |
| Sleep quality (ref very good/good) |  |  |  |  |
|  | Average | 40.71% | 41.26% | 28.36% |
|  | Not good/very poor | 46.33% | 49.04% | 62.21% |
| Smoking (ref non-smoker) |  | 8.41% | 13.26% | 13.43% |
| More than 14 weekly alcoholic drinks (ref =< 14) |  | 8.84% | 7.44% | 8.36% |
| Number of mental health care behaviours (range 0-9) |  | 0.85 (1.26) | 0.80 (1.11) | 0.93 (1.12) |
| **COVID-19 infection variables** |  |  |  |  |
| COVID-19 infection severity in first two weeks (ref asymptomatic) |  |  |  |  |
|  | Mild | 18.12% | 16.68% | 3.96% |
|  | Moderate | 65.58% | 71.58% | 74.85% |
|  | Severe | 11.83% | 8.53% | 13.99% |
| Dominant strain in the UK at time of COVID-19 infection (ref original variant) |  |  |  |  |
|  | Alpha (1 November 2020 to 30 June 2021) | 47.39% | 51.48% | 39.89% |
|  | Delta (1 July to 30 November 2021) | 35.03% | 37.98% | 40.67% |
|  | Omicron (1 December 2021-) | 0.00% | 0.00% | 0.00% |
| **Socio-demographics** |  |  |  |  |
| Female (ref male) |  | 39.48% | 51.61% | 46.72% |
| Age (ref 60+) |  |  |  |  |
|  | 45-59 | 31.89% | 36.73% | 32.44% |
|  | 30-44 | 12.36% | 22.66% | 3.45% |
|  | 18-29 | 1.21% | 0.62% | 0.00% |
| Ethnic minority groups (ref White) |  | 4.91% | 14.75% | 8.97% |
| Education (ref degree or higher) |  |  |  |  |
|  | A-levels or vocational | 26.03% | 26.08% | 30.11% |
|  | Up to GCSE | 51.14% | 50.58% | 50.34% |
| Low household income (<£30,000) |  | 63.72% | 47.78% | 75.89% |
| Employed (ref not employed) |  | 50.71% | 66.19% | 27.54% |
| Key worker |  | 16.03% | 27.71% | 11.80% |
| Crowded household |  | 13.65% | 15.32% | 14.30% |
| Living arrangement (ref alone) |  |  |  |  |
|  | With others, not with children | 52.90% | 49.62% | 50.55% |
|  | With others, including children | 24.68% | 33.01% | 26.26% |
| Live in a rural area (ref urban) |  | 23.19% | 23.12% | 24.38% |
| **Pre-existing conditions** |  |  |  |  |
| Long-term physical health condition (ref none) |  | 66.34% | 57.66% | 82.25% |
| Long-term mental health condition (ref none) |  | 16.80% | 18.78% | 21.89% |

Note. Data were weighted to the proportions of gender, age, ethnicity, country, and education obtained from the Office for National Statistics. GCSE refers to General Certificate of Secondary Education.

Descriptive statistics for health behaviours in the short vs long COVID groups are shown in Table 1 and results from binary logistic regressions are shown in Table 3. There was an increasing likelihood of long COVID for those with poorer quality sleep (Table 4). In the fully adjusted model, compared to people who had had very good or good quality sleep prior to infection, those who reported average and not good or very poor sleep were 2.4-3.5 times as likely to have developed long COVID (average sleep: odds ratio [OR]: 2.44; 95% confidence interval [CI]: 1.44 to 4.12; not good/very poor sleep: OR: 3.53; 95% CI: 2.01 to 6.21). None of the other health behaviours predicted long COVID status.

**Table 3.** Logistic regressions predicting self-reported long COVID from health behaviours in the month prior to COVID-19 infection, weighted (N = 1,581)

|  | Self-reported long COVID |  |  |  |  |  |
| --- | --- | --- | --- | --- | --- | --- |
|  | OR | SE | T | P | 95% CI | 95% CI |
| **Model 1** |  |  |  |  |  |  |
| Exercise frequency (ref no exercise) |  |  |  |  |  |  |
| <30 min to 2 hours | 0.91 | 0.30 | -0.29 | 0.77 | 0.47 | 1.74 |
| 3+ hours | 0.65 | 0.25 | -1.11 | 0.27 | 0.31 | 1.38 |
| Days of fresh air (> 15 minutes) | 0.96 | 0.05 | -0.72 | 0.47 | 0.87 | 1.07 |
| Sleep quality (ref very good/good) |  |  |  |  |  |  |
| Average | **2.39** | **0.59** | **3.54** | **<0.01** | **1.47** | **3.86** |
| Not good/very poor | **4.13** | **1.14** | **5.11** | **<0.01** | **2.40** | **7.11** |
| Smoking (ref non-smoker) | 1.74 | 0.84 | 1.16 | 0.25 | 0.68 | 4.47 |
| More than 14 weekly alcoholic drinks (ref =< 14) | 0.60 | 0.18 | -1.65 | 0.10 | 0.33 | 1.10 |
| Number of mental health care behaviours | 1.03 | 0.09 | 0.39 | 0.70 | 0.88 | 1.22 |
| **Model 2** |  |  |  |  |  |  |
| Exercise frequency (ref no exercise) |  |  |  |  |  |  |
| <30 min to 2 hours | 0.99 | 0.33 | -0.04 | 0.97 | 0.51 | 1.91 |
| 3+ hours | 0.71 | 0.28 | -0.87 | 0.39 | 0.33 | 1.54 |
| Days of fresh air (> 15 minutes) | 0.98 | 0.06 | -0.28 | 0.78 | 0.88 | 1.10 |
| Sleep quality (ref very good/good) |  |  |  |  |  |  |
| Average | **2.40** | **0.64** | **3.31** | **<0.01** | **1.43** | **4.04** |
| Not good/very poor | **3.72** | **1.07** | **4.57** | **<0.01** | **2.12** | **6.54** |
| Smoking (ref non-smoker) | 2.19 | 1.22 | 1.40 | 0.16 | 0.73 | 6.54 |
| More than 14 weekly alcoholic drinks (ref =< 14) | 0.70 | 0.24 | -1.02 | 0.31 | 0.36 | 1.38 |
| Number of mental health care behaviours | 0.97 | 0.08 | -0.35 | 0.72 | 0.82 | 1.14 |
| **Model 3** |  |  |  |  |  |  |
| Exercise frequency (ref no exercise) |  |  |  |  |  |  |
| <30 min to 2 hours | 1.02 | 0.36 | 0.06 | 0.95 | 0.51 | 2.05 |
| 3+ hours | 0.62 | 0.24 | -1.22 | 0.22 | 0.28 | 1.34 |
| Days of fresh air (> 15 minutes) | 1.00 | 0.06 | -0.05 | 0.96 | 0.89 | 1.12 |
| Sleep quality (ref very good/good) |  |  |  |  |  |  |
| Average | **2.43** | **0.64** | **3.35** | **<0.01** | **1.44** | **4.08** |
| Not good/very poor | **3.54** | **1.02** | **4.41** | **<0.01** | **2.02** | **6.21** |
| Smoking (ref non-smoker) | 1.71 | 0.66 | 1.41 | 0.16 | 0.81 | 3.62 |
| More than 14 weekly alcoholic drinks (ref =< 14) | 0.72 | 0.27 | -0.86 | 0.39 | 0.35 | 1.51 |
| Number of mental health care behaviours | 1.06 | 0.09 | 0.73 | 0.47 | 0.90 | 1.26 |
| **Model 4** |  |  |  |  |  |  |
| Exercise frequency (ref no exercise) |  |  |  |  |  |  |
| <30 min to 2 hours | 0.95 | 0.34 | -0.13 | 0.89 | 0.48 | 1.91 |
| 3+ hours | 0.57 | 0.23 | -1.39 | 0.16 | 0.26 | 1.26 |
| Days of fresh air (> 15 minutes) | 1.00 | 0.06 | -0.07 | 0.94 | 0.89 | 1.11 |
| Sleep quality (ref very good/good) |  |  |  |  |  |  |
| Average | **2.44** | **0.65** | **3.33** | **<0.01** | **1.44** | **4.12** |
| Not good/very poor | **3.53** | **1.02** | **4.38** | **<0.01** | **2.01** | **6.21** |
| Smoking (ref non-smoker) | 1.72 | 0.64 | 1.47 | 0.14 | 0.83 | 3.57 |
| More than 14 weekly alcoholic drinks (ref =< 14) | 0.73 | 0.27 | -0.83 | 0.40 | 0.35 | 1.53 |
| Number of mental health care behaviours | 1.10 | 0.10 | 1.13 | 0.26 | 0.93 | 1.31 |

Note. Data were weighted to the proportions of gender, age, ethnicity, country, and education obtained from the Office for National Statistics. Model 1 included only health behaviours (in the same model), Model 2 additionally adjusted for COVID-19 infection variables, Model 3 additionally adjusted for socio-demographic characteristics, and Model 4 additionally adjusted for pre-existing conditions.

**Table 4.**
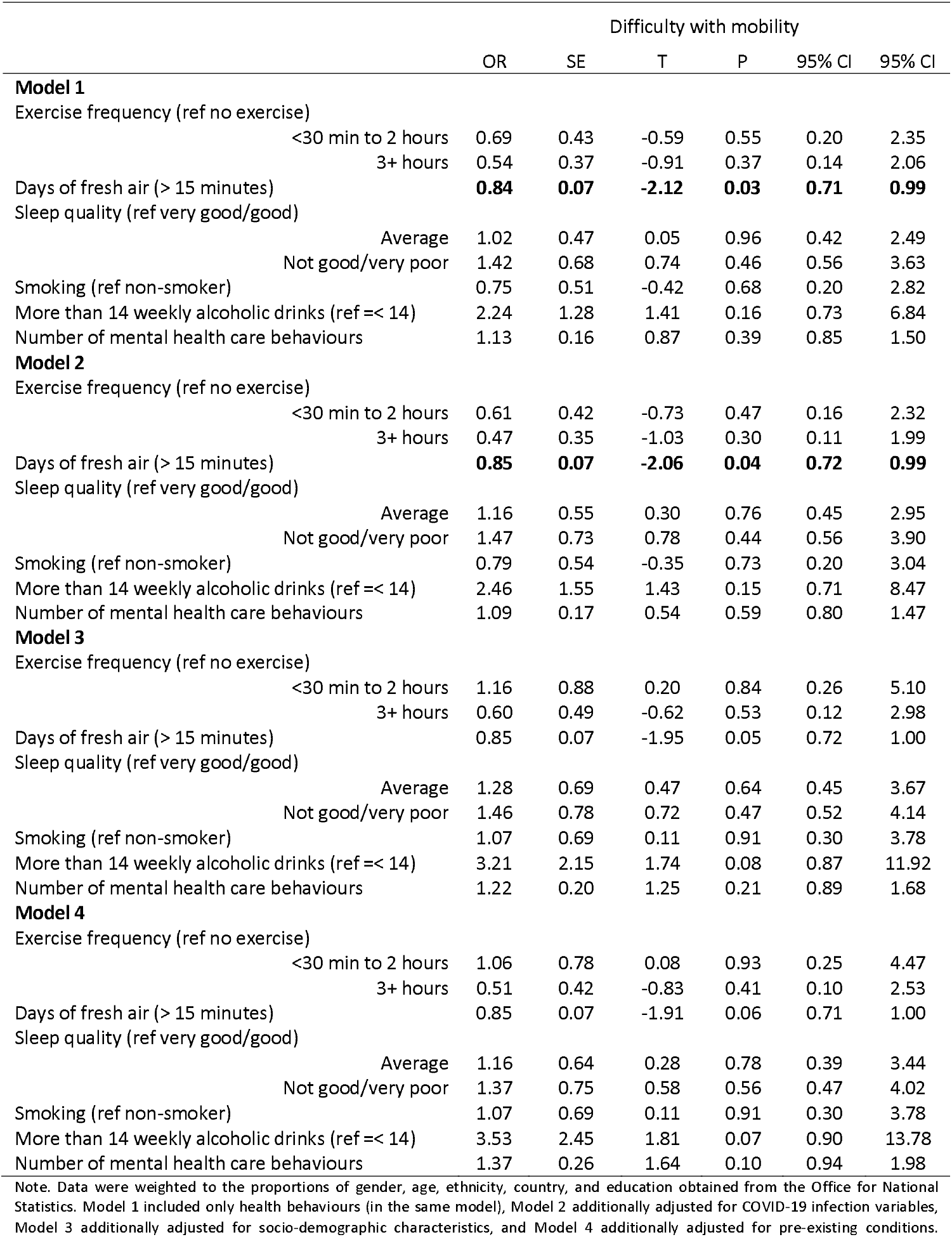
Logistic regressions predicting the development of difficulty with mobility from health behaviours in the month prior to COVID-19 infection, weighted (N = 293)

**Table 5.** Logistic regressions predicting the development of difficulty with cognition from health behaviours in the month prior to COVID-19 infection, weighted (N = 293)

|  | Difficulty with cognition |  |  |  |  |  |
| --- | --- | --- | --- | --- | --- | --- |
|  | OR | SE | T | P | 95% CI | 95% CI |
| **Model 1** |  |  |  |  |  |  |
| Exercise frequency (ref no exercise) |  |  |  |  |  |  |
| <30 min to 2 hours | 1.54 | 0.98 | 0.68 | 0.50 | 0.44 | 5.38 |
| 3+ hours | 1.85 | 1.28 | 0.88 | 0.38 | 0.47 | 7.20 |
| Days of fresh air (> 15 minutes) | 0.92 | 0.09 | -0.85 | 0.40 | 0.77 | 1.11 |
| Sleep quality (ref very good/good) |  |  |  |  |  |  |
| Average | 1.27 | 0.59 | 0.52 | 0.60 | 0.51 | 3.15 |
| Not good/very poor | **3.16** | **1.63** | **2.23** | **0.03** | **1.15** | **8.69** |
| Smoking (ref non-smoker) | 3.27 | 2.68 | 1.45 | 0.15 | 0.66 | 16.30 |
| More than 14 weekly alcoholic drinks (ref =< 14) | 1.26 | 0.66 | 0.44 | 0.66 | 0.45 | 3.54 |
| Number of mental health care behaviours | 1.08 | 0.26 | 0.31 | 0.76 | 0.67 | 1.72 |
| **Model 2** |  |  |  |  |  |  |
| Exercise frequency (ref no exercise) |  |  |  |  |  |  |
| <30 min to 2 hours | 1.18 | 0.79 | 0.25 | 0.80 | 0.32 | 4.36 |
| 3+ hours | 1.36 | 0.99 | 0.42 | 0.67 | 0.32 | 5.68 |
| Days of fresh air (> 15 minutes) | 0.92 | 0.08 | -0.99 | 0.32 | 0.78 | 1.08 |
| Sleep quality (ref very good/good) |  |  |  |  |  |  |
| Average | 1.20 | 0.53 | 0.40 | 0.69 | 0.50 | 2.84 |
| Not good/very poor | 2.63 | 1.32 | 1.92 | 0.05 | 0.98 | 7.03 |
| Smoking (ref non-smoker) | 4.03 | 4.14 | 1.36 | 0.18 | 0.54 | 30.18 |
| More than 14 weekly alcoholic drinks (ref =< 14) | 1.68 | 0.90 | 0.97 | 0.33 | 0.59 | 4.81 |
| Number of mental health care behaviours | 1.01 | 0.26 | 0.03 | 0.97 | 0.61 | 1.67 |
| **Model 3** |  |  |  |  |  |  |
| Exercise frequency (ref no exercise) |  |  |  |  |  |  |
| <30 min to 2 hours | 0.94 | 0.79 | -0.07 | 0.95 | 0.18 | 4.87 |
| 3+ hours | 0.88 | 0.78 | -0.14 | 0.89 | 0.16 | 4.98 |
| Days of fresh air (> 15 minutes) | 0.91 | 0.08 | -1.06 | 0.29 | 0.76 | 1.09 |
| Sleep quality (ref very good/good) |  |  |  |  |  |  |
| Average | 1.05 | 0.57 | 0.09 | 0.93 | 0.36 | 3.06 |
| Not good/very poor | 3.09 | 1.81 | 1.92 | 0.05 | 0.98 | 9.76 |
| Smoking (ref non-smoker) | 2.50 | 1.96 | 1.17 | 0.24 | 0.54 | 11.65 |
| More than 14 weekly alcoholic drinks (ref =< 14) | 1.41 | 0.97 | 0.50 | 0.62 | 0.37 | 5.45 |
| Number of mental health care behaviours | 1.17 | 0.31 | 0.61 | 0.54 | 0.70 | 1.97 |
| **Model 4** |  |  |  |  |  |  |
| Exercise frequency (ref no exercise) |  |  |  |  |  |  |
| <30 min to 2 hours | 0.94 | 0.78 | -0.08 | 0.94 | 0.18 | 4.80 |
| 3+ hours | 0.87 | 0.77 | -0.16 | 0.88 | 0.15 | 4.92 |
| Days of fresh air (> 15 minutes) | 0.90 | 0.08 | -1.10 | 0.27 | 0.76 | 1.08 |
| Sleep quality (ref very good/good) |  |  |  |  |  |  |
| Average | 1.02 | 0.57 | 0.04 | 0.97 | 0.35 | 3.02 |
| Not good/very poor | 3.06 | 1.81 | 1.89 | 0.06 | 0.96 | 9.74 |
| Smoking (ref non-smoker) | 2.46 | 1.91 | 1.16 | 0.25 | 0.54 | 11.30 |
| More than 14 weekly alcoholic drinks (ref =< 14) | 1.36 | 0.94 | 0.45 | 0.65 | 0.35 | 5.26 |
| Number of mental health care behaviours | 1.17 | 0.32 | 0.58 | 0.56 | 0.69 | 2.00 |

Note. Data were weighted to the proportions of gender, age, ethnicity, country, and education obtained from the Office for National Statistics. Model 1 included only health behaviours (in the same model), Model 2 additionally adjusted for COVID-19 infection variables, Model 3 additionally adjusted for socio-demographic characteristics, and Model 4 additionally adjusted for pre-existing conditions.

For individuals with long COVID, descriptive statistics for health behaviours by different long COVID symptoms are shown in Table 2. More days of fresh air for at least 15 minutes reduced the likelihood of difficulty with mobility (OR: 0.85; 95% CI: 0.72 to 0.99; Table 4) but this association was not maintained once socio-demographics and pre-existing health conditions were accounted for (OR: 0.85; 95% CI: 0.71 to 1.00). Not good or very poor-quality sleep associated with increased likelihood of difficulty with cognition (OR: 3.16; 95% CI: 1.15 to 8.69) but not once covariates were included in the model (OR: 3.06; 95% CI: 0.96 to 9.74). Having smoked in the month prior to infection with COVID-19 resulted in a more than eight-fold increased risk (OR: 8.39; 95% CI: 1.86 to 37.91; Table 6) of having difficulty with self-care, whilst weekly exercise of at least three hours reduced this likelihood (OR: 0.05; 95% CI: 0.01 to 0.39) in the fully adjusted model.

**Table 6.** Logistic regressions predicting the development of difficulty with self-care from health behaviours in the month prior to COVID-19 infection, weighted (N = 293)

|  | Difficulty with self-care |  |  |  |  |  |
| --- | --- | --- | --- | --- | --- | --- |
|  | OR | SE | T | P | 95% CI | 95% CI |
| **Model 1** |  |  |  |  |  |  |
| Exercise frequency (ref no exercise) |  |  |  |  |  |  |
| <30 min to 2 hours | 0.82 | 0.58 | -0.28 | 0.78 | 0.20 | 3.30 |
| 3+ hours | 0.27 | 0.26 | -1.33 | 0.18 | 0.04 | 1.85 |
| Days of fresh air (> 15 minutes) | 0.83 | 0.08 | -1.89 | 0.06 | 0.68 | 1.01 |
| Sleep quality (ref very good/good) |  |  |  |  |  |  |
| Average | 0.94 | 0.65 | -0.09 | 0.93 | 0.24 | 3.66 |
| Not good/very poor | 2.36 | 1.56 | 1.29 | 0.20 | 0.64 | 8.63 |
| Smoking (ref non-smoker) | 1.75 | 0.98 | 1.00 | 0.32 | 0.58 | 5.26 |
| More than 14 weekly alcoholic drinks (ref =< 14) | 1.39 | 0.85 | 0.55 | 0.59 | 0.42 | 4.61 |
| Number of mental health care behaviours | 0.97 | 0.17 | -0.14 | 0.89 | 0.69 | 1.38 |
| **Model 2** |  |  |  |  |  |  |
| Exercise frequency (ref no exercise) |  |  |  |  |  |  |
| <30 min to 2 hours | 0.78 | 0.64 | -0.30 | 0.76 | 0.16 | 3.84 |
| 3+ hours | 0.25 | 0.28 | -1.22 | 0.22 | 0.03 | 2.31 |
| Days of fresh air (> 15 minutes) | 0.82 | 0.09 | -1.73 | 0.08 | 0.66 | 1.03 |
| Sleep quality (ref very good/good) |  |  |  |  |  |  |
| Average | 0.93 | 0.75 | -0.08 | 0.93 | 0.19 | 4.49 |
| Not good/very poor | 2.02 | 1.59 | 0.89 | 0.37 | 0.43 | 9.45 |
| Smoking (ref non-smoker) | 1.83 | 1.23 | 0.90 | 0.37 | 0.49 | 6.83 |
| More than 14 weekly alcoholic drinks (ref =< 14) | 1.70 | 1.20 | 0.75 | 0.45 | 0.43 | 6.80 |
| Number of mental health care behaviours | 0.89 | 0.16 | -0.66 | 0.51 | 0.62 | 1.27 |
| **Model 3** |  |  |  |  |  |  |
| Exercise frequency (ref no exercise) |  |  |  |  |  |  |
| <30 min to 2 hours | 0.99 | 0.79 | -0.01 | 0.99 | 0.21 | 4.74 |
| 3+ hours | 0.16 | 0.15 | -1.95 | 0.05 | 0.03 | 1.01 |
| Days of fresh air (> 15 minutes) | 0.84 | 0.09 | -1.52 | 0.13 | 0.68 | 1.05 |
| Sleep quality (ref very good/good) |  |  |  |  |  |  |
| Average | 0.54 | 0.45 | -0.74 | 0.46 | 0.11 | 2.73 |
| Not good/very poor | 0.69 | 0.58 | -0.44 | 0.66 | 0.13 | 3.56 |
| Smoking (ref non-smoker) | **3.95** | **2.75** | **1.97** | **0.05** | **1.01** | **15.46** |
| More than 14 weekly alcoholic drinks (ref =< 14) | 1.85 | 1.47 | 0.77 | 0.44 | 0.39 | 8.80 |
| Number of mental health care behaviours | 0.78 | 0.16 | -1.17 | 0.24 | 0.52 | 1.18 |
| **Model 4** |  |  |  |  |  |  |
| Exercise frequency (ref no exercise) |  |  |  |  |  |  |
| <30 min to 2 hours | 0.66 | 0.48 | -0.56 | 0.57 | 0.16 | 2.78 |
| 3+ hours | **0.05** | **0.05** | **-2.83** | **<0.01** | **0.01** | **0.39** |
| Days of fresh air (> 15 minutes) | 0.80 | 0.10 | -1.85 | 0.06 | 0.63 | 1.01 |
| Sleep quality (ref very good/good) |  |  |  |  |  |  |
| Average | 0.41 | 0.37 | -0.99 | 0.32 | 0.07 | 2.36 |
| Not good/very poor | 0.71 | 0.66 | -0.37 | 0.71 | 0.12 | 4.35 |
| Smoking (ref non-smoker) | **8.39** | **6.45** | **2.76** | **0.01** | **1.86** | **37.91** |
| More than 14 weekly alcoholic drinks (ref =< 14) | 1.01 | 0.81 | 0.01 | 0.99 | 0.21 | 4.88 |
| Number of mental health care behaviours | 0.86 | 0.23 | -0.57 | 0.57 | 0.50 | 1.46 |

Note. Data were weighted to the proportions of gender, age, ethnicity, country, and education obtained from the Office for National Statistics. Model 1 included only health behaviours (in the same model), Model 2 additionally adjusted for COVID-19 infection variables, Model 3 additionally adjusted for socio-demographic characteristics, and Model 4 additionally adjusted for pre-existing conditions.

Results from sensitivity analyses which included people who were unsure whether they had had long COVID indicated similar findings, but with some minor exceptions (Supplemental Tables S10–S13). Having smoked in the month prior to infection with COVID-19 increased the likelihood of long COVID (OR: 1.90; 95% CI: 1.05 to 3.43). Having engaged in more mental health care behaviours predicted difficulty with mobility amongst those with long COVID (OR: 1.38; 95% CI: 1.03 to 1.85; Supplemental Table S11), whilst consumption of more than 14 alcoholic drinks in a single week in the month before infection with COVID-19 increased the likelihood of difficulty with self-care by 5.24 (95% CI: 1.34 to 19.58; Supplemental Table S13). Being slightly or very overweight was associated with 1.63 times greater odds (95% CI: 1.04 to 2.55) of long COVID (Supplemental Table S14). Findings for individual long COVID symptoms were different from those in the main analyses when overweight status was included. Not good/very poor-quality sleep and more mental health care behaviours predicted difficulty with cognition (Supplemental Table S16), and none of the health behaviours associated with the other long COVID symptoms.

## Discussion

This study explored the relationship between modifiable health behaviours in the month preceding COVID-19 infection and the risk of developing long COVID in a longitudinal study of UK adults. Notably, there was little evidence of associations, with no relationship found in unadjusted or adjusted models in physical activity, fresh air, smoking, alcohol consumption or mental health care behaviours. The only association that was found was with sleep, with poorer sleep in the month prior to infection associated with a 2.4-3.5-fold increase in risk of developing long COVID. Similarly, amongst participants who did develop long COVID, there was little evidence of association between health behaviours and mobility or cognition difficulties. But regular daily exercise was associated with 95% lower odds of developing difficulties with self-care, whilst smoking was associated with more than an 8-fold increase in risk of developing such difficulties.

Comparison with other research on pre-infection health behaviours is difficult due to their being a lack of research in this area. Research on pre-infection factors influencing long COVID has focused on health conditions such as obesity, asthma, and higher pre-pandemic levels of psychological distress,^4,7–10^ but research on health behaviours, which may be more amenable to intervention, is largely absent. Several of the health behaviours we examined, such as not smoking, a healthy diet, regular physical activity, and consuming fewer than 14 alcoholic drinks per week are all inversely associated with all-cause mortality and longer lifespan.^27^ Nevertheless, our null findings are congruent with other research showing that identifying who is most at risk for long COVID is difficult to determine, even with factors such as symptom severity sometimes showing inconsistent associations with long COVID development.^6,28–30^

However, we did find associations between poor sleep and development of long COVID. Whilst sleep disturbances have been commonly reported in people suffering from long COVID,^31,32^ to our knowledge this is the first study to examine pre-infection sleep quality in relation to long COVID development. Lack of sleep can compromise both innate and adaptive immune function, making individuals more susceptible to infectious disease, and reduce the effectiveness of vaccine response.^14,15^ Therefore, it is possible that by depleting physical resources, poor sleep places the body in a more vulnerable state for tackling COVID-19 infection. However, it is also possible that poor sleep is an indicator of other psychological stressors that could in fact be the cause of a heightened risk of developing long COVID. Prior work from our research group, which also used data from the UCL COVID-19 Social Study, reported elevated risk from worrying about a range of adversity experiences in the month before becoming infected with COVID-19 and developing long COVID.^33^ Experiencing adversities as well as worrying about adversities also predicted lower sleep quality in the first months of the pandemic,^34^ suggesting a potential biobehavioural pathway from stress to long COVID via impaired sleep.^35^

Amongst individuals who did develop long COVID, we also found an association between exercise and the risk of developing problems involving self-care. Meeting weekly physical activity guidelines (at least 3 hours a week) was associated with a reduced odds of self-care difficulties, but this was not found for less frequent activity (e.g., 30 mins – 2 hours a week). Our findings echo those from a study that focused on physical activity pre-pandemic and risk of adverse COVID-19 outcomes including hospitalisation, mortality, and the need for intensive care in a large US study.^36^ But our results extend these by showing that physical activity remains important in the weeks immediately prior to infection. Regular physical activity plays a critical role in reducing risk for acquiring and death from infectious disease, strengthening the immune system, and enhancing vaccine response.^13^ Exercise may therefore attenuate COVID-19 sequelae and persistent symptoms by moderating the inflammatory response.^37^ Despite these potential benefits, the relationship between exercise and long COVID symptoms appears to be complex. Although exercise may improve symptoms of long COVID, long COVID symptoms can be triggered by physical activity,^38^ and should therefore be titrated according to individual patient needs.^1,39^ Therefore, it is important that in potential future pandemics, public health guidelines include an emphasis on maintaining physical activity, as this could help to reduce the functional consequences of developing long COVID.

Finally, having smoked in the month prior to becoming infected with COVID-19 was by far the largest predictor of difficulties with self-care amongst adults with long COVID. Although smoking has been associated with increased likelihood of more severe COVID-19 outcomes including mortality,^40^ to our knowledge only one other study has examined smoking as a risk factor for long COVID. Findings from a sample designed to be representative of the adult population in England, showed that the odds of persistent COVID-19 symptoms (12 weeks or more) from smoking (OR: 1.35) were higher than those of being overweight (OR: 1.16).^4^ However, smoking was measured concurrently with outcomes. Our study therefore extends these findings by showing a temporal relationship between smoking and the development of specific long COVID symptoms. Although the prevalence of smokers in England decreased over the period 2011 to 2019 from 19.8% to 13.9%, certain groups continue to be more likely to smoke: people with a mental health condition and those working in lower skilled occupations.^16^ Lower socio-economic status and pre-existing mental health conditions have both been found to be risk factors for developing long COVID,^4,10,33^ underscoring the importance of smoking cessation particularly for vulnerable groups. Some long COVID management guidelines recommend not smoking to manage symptoms such as breathlessness,^41^ but this advice is currently only within a subsection of the National Health Service’s COVID recovery guidance for patients which advises people to avoid smoking or vaping near their oxygen tank at home.^42^ General advice about not smoking is absent, although information about eating well, physical activity, and sleeping well is provided. Our findings suggest the importance of highlighting the risks of smoking in relation to potential future symptoms of long COVID.

This study has several strengths as well as limitations. A major strength is its longitudinal design, particularly the measurement of health behaviours prior to infection with COVID-19, the latter of which is random and cannot be predicted. However, due to data limitations, we were not able to include important health behaviours such as diet and nutrition, which are key behavioural risks for morbidity.^16^ We also assessed a limited number of long COVID symptoms, and did not assess fatigue, which is often the most commonly reported.^5^ Although well-stratified across major demographic groups, the study sample is also not representative of the general UK population, and results therefore cannot be generalised.

Our findings add to the dearth of research on health behaviours prior to infection with COVID-19 and the development of long COVID and suggest the importance of regular physical activity and smoking cessation, as early intervention (especially in relation to smoking) may reduce the likelihood of long COVID amongst those who become infected. Poor quality sleep prior to infection with COVID-19 is also associated with the development of long COVID but may be a result of stress due to experience of or concerns about adversities. More research on risk factors for long COVID is important, given that it is not the health behaviours per se that cause long COVID. At the time of writing, the percentage of people in the UK testing positive for COVID-19 is at an all-time high and still increasing,^43^ and numbers of the population experiencing long COVID is estimated to be 2.7% of the general population, which means an estimated 1.7 million people are experiencing long COVID symptoms.^3^ With the removal of free testing in the UK, it is important to promote public health messages to help people minimise their risk of developing long-term debilitating symptoms.

## Data Availability

The UCL COVID-19 Social Study documentation and codebook are available for download at https://osf.io/jm8ra/. Statistical code is available upon request from Elise Paul.

## Contributors

DF conceptualised and designed the study. DF also acquired funding, led the investigation, provided oversight on the methodology, administered the project, provided software and other resources, and supervised the project. Data were curated, validated, and formally analysed by EP. EP created visualisations, wrote the original manuscript draft with input from all authors. All authors reviewed and edited the manuscript. All authors approved the final version of the manuscript and had full access to and verified the data.

## Declaration of interests

All authors declare no conflicts of interest.

## Acknowledgements

The researchers are grateful for the support of a number of organisations with their recruitment efforts including: the UKRI Mental Health Networks, Find Out Now, UCL BioResource, SEO Works, FieldworkHub, and Optimal Workshop.

## Ethics approval and consent to participate

Ethical approval for the COVID-19 Social Study was granted by the UCL Ethics Committee. All participants provided fully informed consent and the study is GDPR compliant.

## Supplemental Materials

**Table S1.** Pattern of missing data in study sample (N = 1,811)

| Variable | Proportion with missing data |
| --- | --- |
| Household income | 8.01% |
| Employment status | 0 |
| Key worker status | 0 |
| Household crowding status | 0 |
| Living arrangement | 0 |
| Live in a rural area | 0 |
| COVID-19 infection severity | 0.83% |
| Dominant strain in the UK at time of COVID-19 infection | 0 |
| Long-term physical health condition | 0 |
| Long-term mental health condition | 0 |
| Exercise frequency | 0.11% |
| Number of days of fresh air | 0.44% |
| Sleep quality | 0 |
| Smoking | 0 |
| More than 14 weekly alcoholic drinks (ref =< 14) consumed in a single week | 0 |
| Number of mental health care behaviours | 0.66% |

**Table S2.**
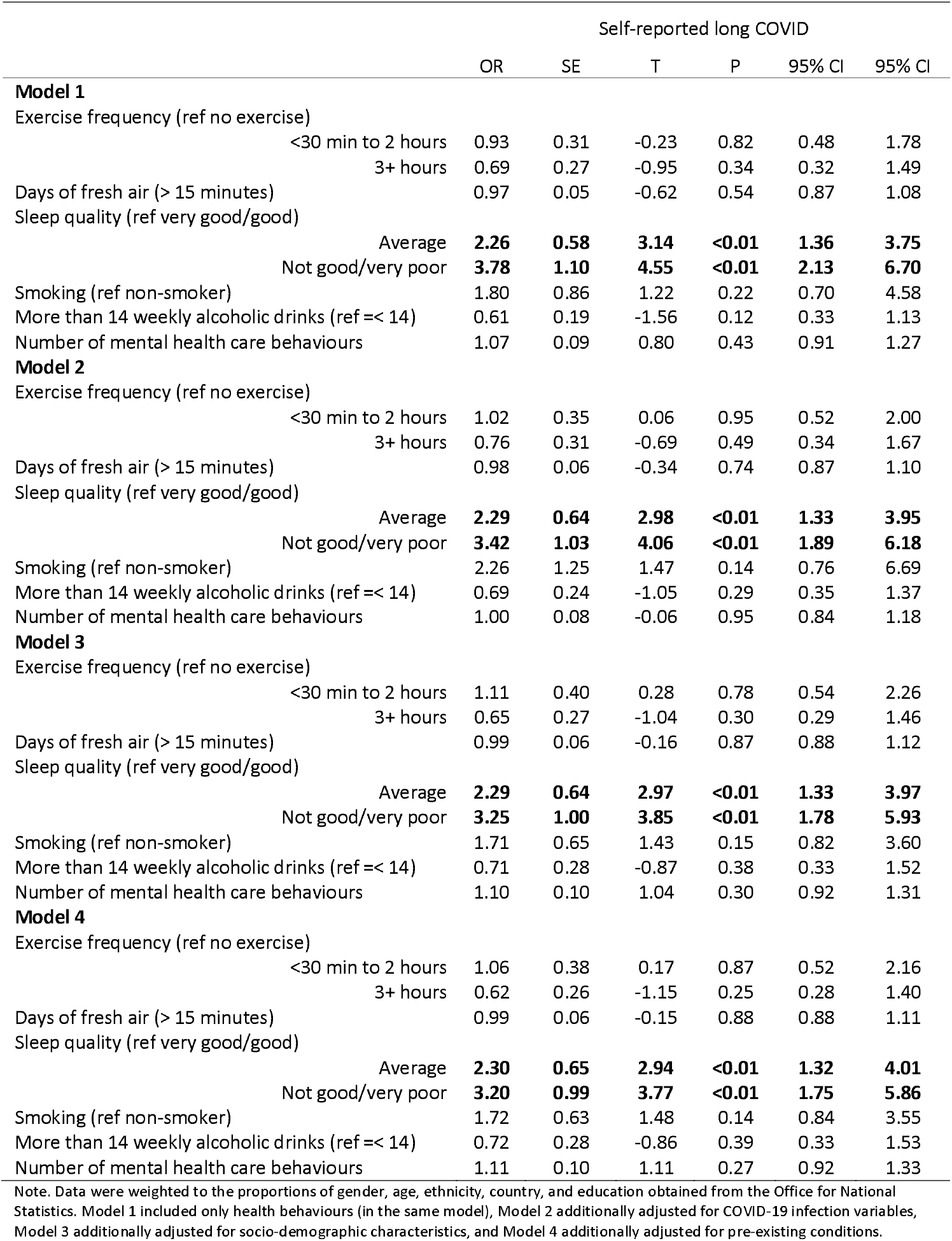
Complete case analysis: logistic regressions predicting the development of long COVID from health behaviours in the month prior to COVID-19 infection, weighted (N = 1,430)

**Table S3.** Complete case analysis: logistic regressions predicting the development of difficulty with mobility from health behaviours in the month prior to COVID-19 infection, weighted (N =264)

|  |  | Difficulty with mobility |  |  |  |  |  |
| --- | --- | --- | --- | --- | --- | --- | --- |
|  |  | OR | SE | T | P | 95% CI | 95% CI |
| **Model 1** |  |  |  |  |  |  |  |
| Exercise frequency (ref no exercise) |  |  |  |  |  |  |  |
|  | <30 min to 2 hours | 0.60 | 0.38 | -0.80 | 0.42 | 0.17 | 2.10 |
|  | 3+ hours | 0.51 | 0.37 | -0.93 | 0.35 | 0.13 | 2.08 |
| Days of fresh air (> 15 minutes) |  | 0.85 | 0.07 | -1.82 | 0.07 | 0.72 | 1.01 |
| Sleep quality (ref very good/good) |  |  |  |  |  |  |  |
|  | Average | 0.89 | 0.42 | -0.24 | 0.81 | 0.35 | 2.26 |
|  | Not good/very poor | 1.17 | 0.59 | 0.31 | 0.76 | 0.44 | 3.13 |
| Smoking |  | 0.76 | 0.52 | -0.40 | 0.69 | 0.20 | 2.88 |
| More than 14 weekly alcoholic drinks (ref =< 14) |  | 2.51 | 1.51 | 1.52 | 0.13 | 0.77 | 8.16 |
| Number of mental health care behaviours |  | 1.24 | 0.19 | 1.43 | 0.15 | 0.92 | 1.68 |
| **Model 2** |  |  |  |  |  |  |  |
| Exercise frequency (ref no exercise) |  |  |  |  |  |  |  |
|  | <30 min to 2 hours | 0.48 | 0.35 | -0.99 | 0.32 | 0.11 | 2.03 |
|  | 3+ hours | 0.45 | 0.35 | -1.02 | 0.31 | 0.10 | 2.10 |
| Days of fresh air (> 15 minutes) |  | 0.87 | 0.08 | -1.61 | 0.11 | 0.73 | 1.03 |
| Sleep quality (ref very good/good) |  |  |  |  |  |  |  |
|  | Average | 1.10 | 0.54 | 0.19 | 0.85 | 0.42 | 2.89 |
|  | Not good/very poor | 1.26 | 0.66 | 0.44 | 0.66 | 0.45 | 3.49 |
| Smoking (ref non-smoker) |  | 0.77 | 0.53 | -0.38 | 0.71 | 0.20 | 2.95 |
| More than 14 weekly alcoholic drinks (ref =< 14) |  | 2.93 | 1.86 | 1.69 | 0.09 | 0.84 | 10.16 |
| Number of mental health care behaviours |  | 1.20 | 0.19 | 1.12 | 0.26 | 0.87 | 1.64 |
| **Model 3** |  |  |  |  |  |  |  |
| Exercise frequency (ref no exercise) |  |  |  |  |  |  |  |
|  | <30 min to 2 hours | 1.08 | 0.86 | 0.10 | 0.92 | 0.23 | 5.11 |
|  | 3+ hours | 0.63 | 0.53 | -0.55 | 0.58 | 0.12 | 3.24 |
| Days of fresh air (> 15 minutes) |  | 0.86 | 0.08 | -1.70 | 0.09 | 0.72 | 1.02 |
| Sleep quality (ref very good/good) |  |  |  |  |  |  |  |
|  | Average | 1.23 | 0.68 | 0.38 | 0.71 | 0.42 | 3.62 |
|  | Not good/very poor | 1.40 | 0.76 | 0.62 | 0.53 | 0.48 | 4.07 |
| Smoking (ref non-smoker) |  | 1.12 | 0.69 | 0.18 | 0.85 | 0.33 | 3.77 |
| More than 14 weekly alcoholic drinks (ref =< 14) |  | **3.92** | **2.71** | **1.97** | **0.05** | **1.01** | **15.22** |
| Number of mental health care behaviours |  | 1.26 | 0.21 | 1.38 | 0.17 | 0.91 | 1.76 |
| **Model 4** |  |  |  |  |  |  |  |
| Exercise frequency (ref no exercise) |  |  |  |  |  |  |  |
|  | <30 min to 2 hours | 0.94 | 0.70 | -0.08 | 0.94 | 0.22 | 4.06 |
|  | 3+ hours | 0.52 | 0.43 | -0.78 | 0.43 | 0.10 | 2.66 |
| Days of fresh air (> 15 minutes) |  | 0.86 | 0.08 | -1.66 | 0.10 | 0.71 | 1.03 |
| Sleep quality (ref very good/good) |  |  |  |  |  |  |  |
|  | Average | 1.19 | 0.67 | 0.31 | 0.76 | 0.39 | 3.59 |
|  | Not good/very poor | 1.39 | 0.78 | 0.59 | 0.55 | 0.46 | 4.18 |
| Smoking (ref non-smoker) |  | 1.12 | 0.72 | 0.18 | 0.85 | 0.32 | 3.93 |
| More than 14 weekly alcoholic drinks (ref =< 14) |  | **4.58** | **3.32** | **2.09** | **0.04** | **1.10** | **19.00** |
| Number of mental health care behaviours |  | 1.43 | 0.28 | 1.80 | 0.07 | 0.97 | 2.11 |

Note. Data were weighted to the proportions of gender, age, ethnicity, country, and education obtained from the Office for National Statistics. Model 1 included only health behaviours (in the same model), Model 2 additionally adjusted for COVID-19 infection variables, Model 3 additionally adjusted for socio-demographic characteristics, and Model 4 additionally adjusted for pre-existing conditions.

**Table S4.** Complete case analysis: logistic regressions predicting the development of difficulty with cognition from health behaviours in the month prior to COVID-19 infection, weighted (N = 264)

|  | Difficulty with cognition |  |  |  |  |  |
| --- | --- | --- | --- | --- | --- | --- |
|  | OR | SE | T | P | 95% CI | 95% CI |
| **Model 1** |  |  |  |  |  |  |
| Exercise frequency (ref no exercise) |  |  |  |  |  |  |
| <30 min to 2 hours | 1.31 | 0.79 | 0.45 | 0.65 | 0.40 | 4.27 |
| 3+ hours | 1.80 | 1.25 | 0.84 | 0.40 | 0.46 | 7.03 |
| Days of fresh air (> 15 minutes) | 0.89 | 0.08 | -1.26 | 0.21 | 0.74 | 1.07 |
| Sleep quality (ref very good/good) |  |  |  |  |  |  |
| Average | 1.21 | 0.59 | 0.39 | 0.70 | 0.47 | 3.13 |
| Not good/very poor | 3.24 | 1.72 | 2.21 | 0.03 | 1.14 | 9.16 |
| Smoking (ref non-smoker) | 3.28 | 2.71 | 1.43 | 0.15 | 0.65 | 16.61 |
| More than 14 weekly alcoholic drinks (ref =< 14) | 1.13 | 0.61 | 0.22 | 0.83 | 0.39 | 3.26 |
| Number of mental health care behaviours | 1.06 | 0.26 | 0.23 | 0.82 | 0.65 | 1.71 |
| **Model 2** |  |  |  |  |  |  |
| Exercise frequency (ref no exercise) |  |  |  |  |  |  |
| <30 min to 2 hours | 1.08 | 0.70 | 0.11 | 0.91 | 0.30 | 3.88 |
| 3+ hours | 1.53 | 1.13 | 0.57 | 0.57 | 0.36 | 6.54 |
| Days of fresh air (> 15 minutes) | 0.89 | 0.08 | -1.33 | 0.18 | 0.74 | 1.06 |
| Sleep quality (ref very good/good) |  |  |  |  |  |  |
| Average | 1.19 | 0.56 | 0.38 | 0.71 | 0.48 | 2.98 |
| Not good/very poor | 2.73 | 1.45 | 1.89 | 0.06 | 0.96 | 7.75 |
| Smoking (ref non-smoker) | 4.15 | 4.29 | 1.38 | 0.17 | 0.55 | 31.47 |
| More than 14 weekly alcoholic drinks (ref =< 14) | 1.59 | 0.89 | 0.83 | 0.41 | 0.53 | 4.77 |
| Number of mental health care behaviours | 0.97 | 0.27 | -0.10 | 0.92 | 0.57 | 1.66 |
| **Model 3** |  |  |  |  |  |  |
| Exercise frequency (ref no exercise) |  |  |  |  |  |  |
| <30 min to 2 hours | 0.98 | 0.75 | -0.03 | 0.98 | 0.22 | 4.37 |
| 3+ hours | 1.15 | 1.00 | 0.16 | 0.87 | 0.21 | 6.35 |
| Days of fresh air (> 15 minutes) | 0.87 | 0.08 | -1.42 | 0.16 | 0.73 | 1.05 |
| Sleep quality (ref very good/good) |  |  |  |  |  |  |
| Average | 1.20 | 0.71 | 0.31 | 0.76 | 0.38 | 3.82 |
| Not good/very poor | **4.23** | **2.79** | **2.19** | **0.03** | **1.17** | **15.38** |
| Smoking (ref non-smoker) | 2.65 | 2.10 | 1.23 | 0.22 | 0.56 | 12.54 |
| More than 14 weekly alcoholic drinks (ref =< 14) | 1.11 | 0.79 | 0.14 | 0.89 | 0.27 | 4.49 |
| Number of mental health care behaviours | 1.09 | 0.28 | 0.35 | 0.73 | 0.66 | 1.00 |
| **Model 4** |  |  |  |  |  |  |
| Exercise frequency (ref no exercise) |  |  |  |  |  |  |
| <30 min to 2 hours | 0.97 | 0.75 | -0.04 | 0.97 | 0.21 | 4.41 |
| 3+ hours | 1.16 | 1.02 | 0.17 | 0.87 | 0.21 | 6.47 |
| Days of fresh air (> 15 minutes) | 0.88 | 0.08 | -1.38 | 0.17 | 0.73 | 1.06 |
| Sleep quality (ref very good/good) |  |  |  |  |  |  |
| Average | 1.24 | 0.73 | 0.37 | 0.71 | 0.39 | 3.92 |
| Not good/very poor | **4.35** | **2.86** | **2.24** | **0.03** | **1.20** | **15.79** |
| Smoking (ref non-smoker) | 2.71 | 2.15 | 1.26 | 0.21 | 0.58 | 12.78 |
| More than 14 weekly alcoholic drinks (ref =< 14) | 1.15 | 0.83 | 0.19 | 0.85 | 0.28 | 4.75 |
| Number of mental health care behaviours | 1.11 | 0.31 | 0.37 | 0.71 | 0.64 | 1.90 |

Note. Data were weighted to the proportions of gender, age, ethnicity, country, and education obtained from the Office for National Statistics. Model 1 included only health behaviours (in the same model), Model 2 additionally adjusted for COVID-19 infection variables, Model 3 additionally adjusted for socio-demographic characteristics, and Model 4 additionally adjusted for pre-existing conditions.

**Table S5.** Complete case analysis: logistic regressions predicting the development of difficulty with self-care from health behaviours in the month prior to COVID-19 infection, weighted (N = 264)

|  |  | Difficulty with self-care |  |  |  |  |  |
| --- | --- | --- | --- | --- | --- | --- | --- |
|  |  | OR | SE | T | P | 95% CI | 95% CI |
| **Model 1** |  |  |  |  |  |  |  |
| Exercise frequency (ref no exercise) |  |  |  |  |  |  |  |
|  | <30 min to 2 hours | 0.61 | 0.41 | -0.73 | 0.46 | 0.17 | 2.25 |
|  | 3+ hours | 0.26 | 0.26 | -1.37 | 0.17 | 0.04 | 1.79 |
| Days of fresh air (> 15 minutes) |  | 0.85 | 0.09 | -1.56 | 0.12 | 0.70 | 1.04 |
| Sleep quality (ref very good/good) |  |  |  |  |  |  |  |
|  | Average | 0.85 | 0.62 | -0.22 | 0.83 | 0.20 | 3.57 |
|  | Not good/very poor | 2.07 | 1.37 | 1.09 | 0.27 | 0.56 | 7.58 |
| Smoking (ref non-smoker) |  | 1.73 | 0.95 | 0.99 | 0.32 | 0.59 | 5.09 |
| More than 14 weekly alcoholic drinks (ref =< 14) |  | 1.41 | 0.86 | 0.57 | 0.57 | 0.43 | 4.64 |
| Number of mental health care behaviours |  | 1.03 | 0.18 | 0.15 | 0.88 | 0.73 | 1.45 |
| **Model 2** |  |  |  |  |  |  |  |
| Exercise frequency (ref no exercise) |  |  |  |  |  |  |  |
|  | <30 min to 2 hours | 0.63 | 0.47 | -0.62 | 0.54 | 0.15 | 2.72 |
|  | 3+ hours | 0.26 | 0.29 | -1.22 | 0.22 | 0.03 | 2.26 |
| Days of fresh air (> 15 minutes) |  | 0.84 | 0.10 | -1.53 | 0.12 | 0.67 | 1.05 |
| Sleep quality (ref very good/good) |  |  |  |  |  |  |  |
|  | Average | 1.00 | 0.83 | 0.00 | 1.00 | 0.20 | 5.09 |
|  | Not good/very poor | 2.13 | 1.72 | 0.94 | 0.35 | 0.44 | 10.38 |
| Smoking (ref non-smoker) |  | 1.89 | 1.25 | 0.96 | 0.34 | 0.52 | 6.89 |
| More than 14 weekly alcoholic drinks (ref =< 14) |  | 1.96 | 1.32 | 1.00 | 0.32 | 0.52 | 7.37 |
| Number of mental health care behaviours |  | 0.88 | 0.16 | -0.70 | 0.48 | 0.62 | 1.25 |
| **Model 3** |  |  |  |  |  |  |  |
| Exercise frequency (ref no exercise) |  |  |  |  |  |  |  |
|  | <30 min to 2 hours | 1.49 | 1.07 | 0.56 | 0.58 | 0.36 | 6.11 |
|  | 3+ hours | **0.18** | **0.15** | **-2.10** | **0.04** | **0.04** | **0.89** |
| Days of fresh air (> 15 minutes) |  | 0.82 | 0.10 | -1.69 | 0.09 | 0.64 | 1.03 |
| Sleep quality (ref very good/good) |  |  |  |  |  |  |  |
|  | Average | 0.59 | 0.45 | -0.69 | 0.49 | 0.13 | 2.63 |
|  | Not good/very poor | 1.26 | 0.97 | 0.30 | 0.76 | 0.28 | 5.71 |
| Smoking (ref non-smoker) |  | **4.76** | **3.09** | **2.40** | **0.02** | **1.33** | **17.01** |
| More than 14 weekly alcoholic drinks (ref =< 14) |  | 1.35 | 1.01 | 0.39 | 0.69 | 0.31 | 5.88 |
| Number of mental health care behaviours |  | 0.72 | 0.17 | -1.38 | 0.17 | 0.45 | 1.15 |
| **Model 4** |  |  |  |  |  |  |  |
| Exercise frequency (ref no exercise) |  |  |  |  |  |  |  |
|  | <30 min to 2 hours | 0.90 | 0.65 | -0.14 | 0.89 | 0.22 | 3.73 |
|  | 3+ hours | **0.07** | **0.07** | **-2.88** | **<0.01** | **0.01** | **0.43** |
| Days of fresh air (> 15 minutes) |  | 0.81 | 0.10 | -1.72 | 0.09 | 0.63 | 1.03 |
| Sleep quality (ref very good/good) |  |  |  |  |  |  |  |
|  | Average | 0.51 | 0.42 | -0.82 | 0.41 | 0.10 | 2.58 |
|  | Not good/very poor | 1.09 | 0.95 | 0.10 | 0.92 | 0.20 | 5.97 |
| Smoking (ref non-smoker) |  | **6.84** | **4.84** | **2.72** | **0.01** | **1.71** | **27.35** |
| More than 14 weekly alcoholic drinks (ref =< 14) |  | 1.07 | 0.87 | 0.08 | 0.93 | 0.22 | 5.28 |
| Number of mental health care behaviours |  | 0.81 | 0.22 | -0.79 | 0.43 | 0.47 | 1.38 |

Note. Data were weighted to the proportions of gender, age, ethnicity, country, and education obtained from the Office for National Statistics. Model 1 included only health behaviours (in the same model), Model 2 additionally adjusted for COVID-19 infection variables, Model 3 additionally adjusted for socio-demographic characteristics, and Model 4 additionally adjusted for pre-existing conditions.

**Table S6.** Characteristics of excluded and included participants, unweighted

|  |  | Excluded<br>N = 30,790<br>Prop. | Included<br>N = 1,811<br>Prop. |
| --- | --- | --- | --- |
| **Long COVID variables** |  |  |  |
| Long COVID status |  |  |  |
|  | Unsure | 16.44% | 12.70% |
|  | Yes, but not formally diagnosed | 12.96% | 12.31% |
|  | Yes, formally diagnosed | 3.54% | 3.87% |
| Presence of specific long COVID symptoms (amongst participants with long COVID) |  |  |  |
|  | Mobility (e.g., walking or climbing steps) | 43.43% | 43.02% |
|  | Cognition (e.g., remembering or concentrating) | 60.54% | 59.27% |
|  | Self-care (e.g., washing all over or dressing) | 17.55% | 10.71% |
| **Health behaviours in month prior to COVID-19 infection** |  |  |  |
| Exercise frequency (ref no exercise) |  |  |  |
|  | <30 min to 2 hours | 56.09% | 67.05% |
|  | 3+ hours | 37.98% | 25.30% |
| Days of fresh air (> 15 minutes) |  | 6.79 (0.77) | 5.78 (1.91) |
| Sleep quality (ref very good/good) |  |  |  |
|  | Average | 20.77% | 41.52% |
|  | Not good/very poor | 72.85% | 24.96% |
| Smoking (ref non-smoker) |  | 9.49% | 5.91% |
| More than 14 weekly alcoholic drinks (ref =< 14) |  | 31.61% | 9.94% |
| Number of mental health care behaviours |  | 1.51 (1.47) | 0.74 (1.05) |
| **COVID-19 infection variables** |  |  |  |
| COVID-19 infection severity in first two weeks (ref asymptomatic) |  |  |  |
|  | Mild | 35.00% | 38.27% |
|  | Moderate | 59.13% | 51.77% |
|  | Severe | 1.94% | 2.40% |
| Dominant strain in the UK at time of COVID-19 infection (ref original variant) |  |  |  |
|  | Alpha (1 November 2020 to 30 June 2021) | 12.48% | 41.58% |
|  | Delta (1 July to 30 November 2021) | 31.93% | 42.19% |
|  | Omicron (1 December 2021-) | 0.47% | 0.00% |
| **Socio-demographics** |  |  |  |
| Female (ref male) |  |  |  |
|  |  | 74.67% | 75.59% |
| Age (ref 60+) |  |  |  |
|  | 45-59 | 41.48% | 40.70% |
|  | 30-44 | 34.16% | 23.91% |
|  | 18-29 | 19.65% | 5.96% |
| Ethnic minority groups (ref White) |  |  |  |
|  |  | 4.71% | 3.64% |
| Education (ref degree or higher) |  |  |  |
|  | A-levels or vocational | 3.76% | 17.01% |
|  | Up to GCSE | 17.18% | 13.09% |
| Low household income (<£30,000) |  |  |  |
|  |  | 14.44% | 34.45% |
| Employed (ref not employed) |  |  |  |
|  |  | 41.30% | 67.59% |
| Key worker |  |  |  |
|  |  | 57.46% | 27.33% |
| Crowded household |  |  |  |
|  |  | 20.92% | 9.28% |
| Living arrangement (ref alone) |  |  |  |
|  | With others, not with children | 8.03% | 52.24% |
|  | With others, including children | 57.27% | 31.36% |
| Live in a rural area (ref urban) |  |  |  |
|  |  | 21.22% | 20.04% |
| **Pre-existing health conditions** |  |  |  |
| Long-term physical health condition (ref none) |  |  |  |
|  |  | 43.56% | 38.87% |
| Long-term mental health condition (ref none) |  |  |  |
|  |  | 16.42% | 15.24% |

**Table S7.**
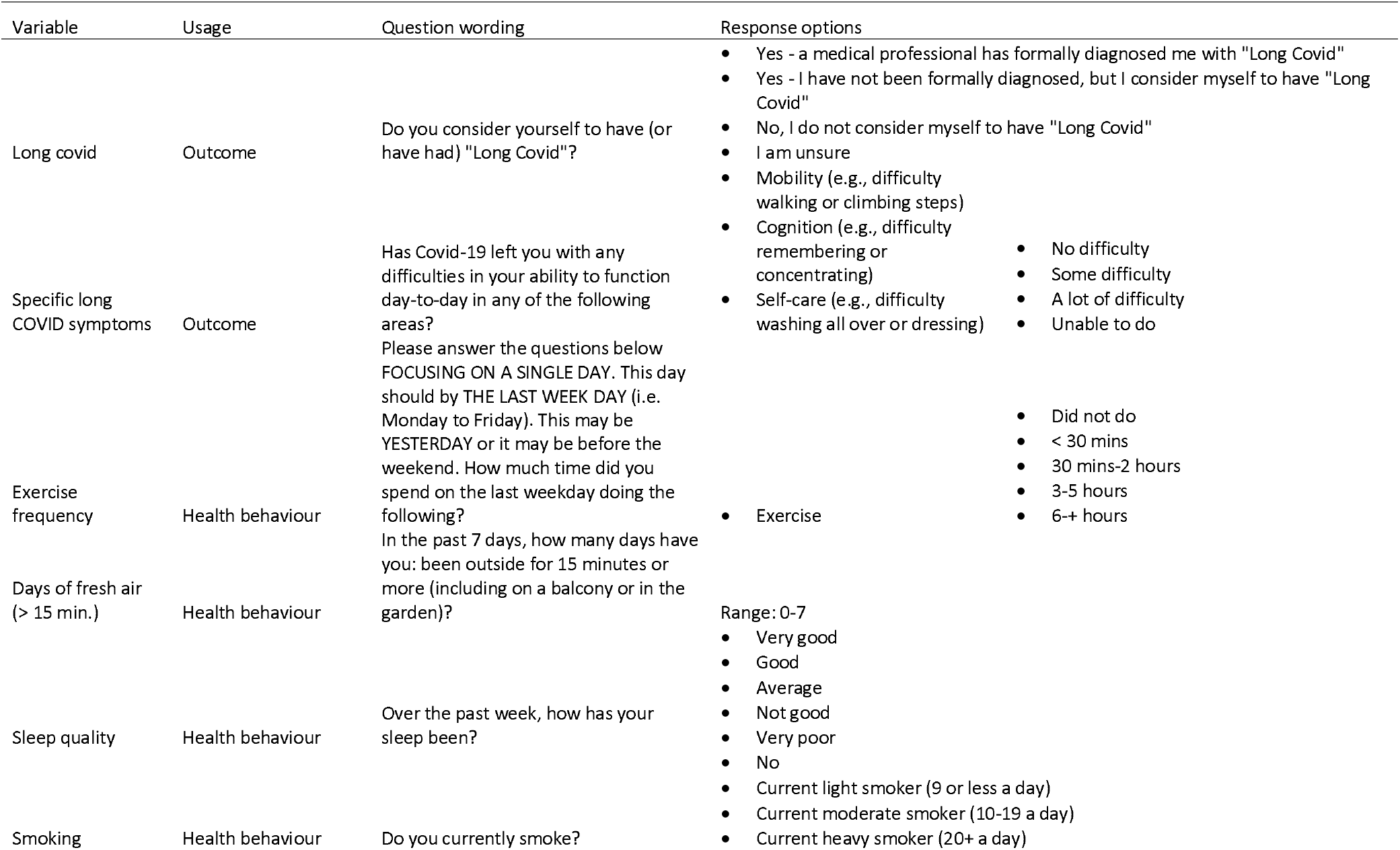
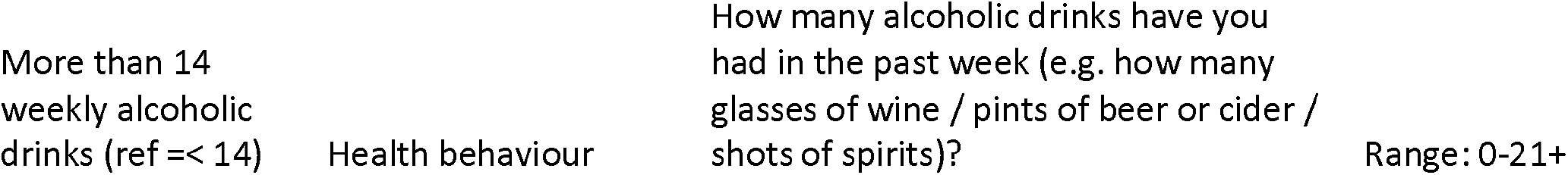
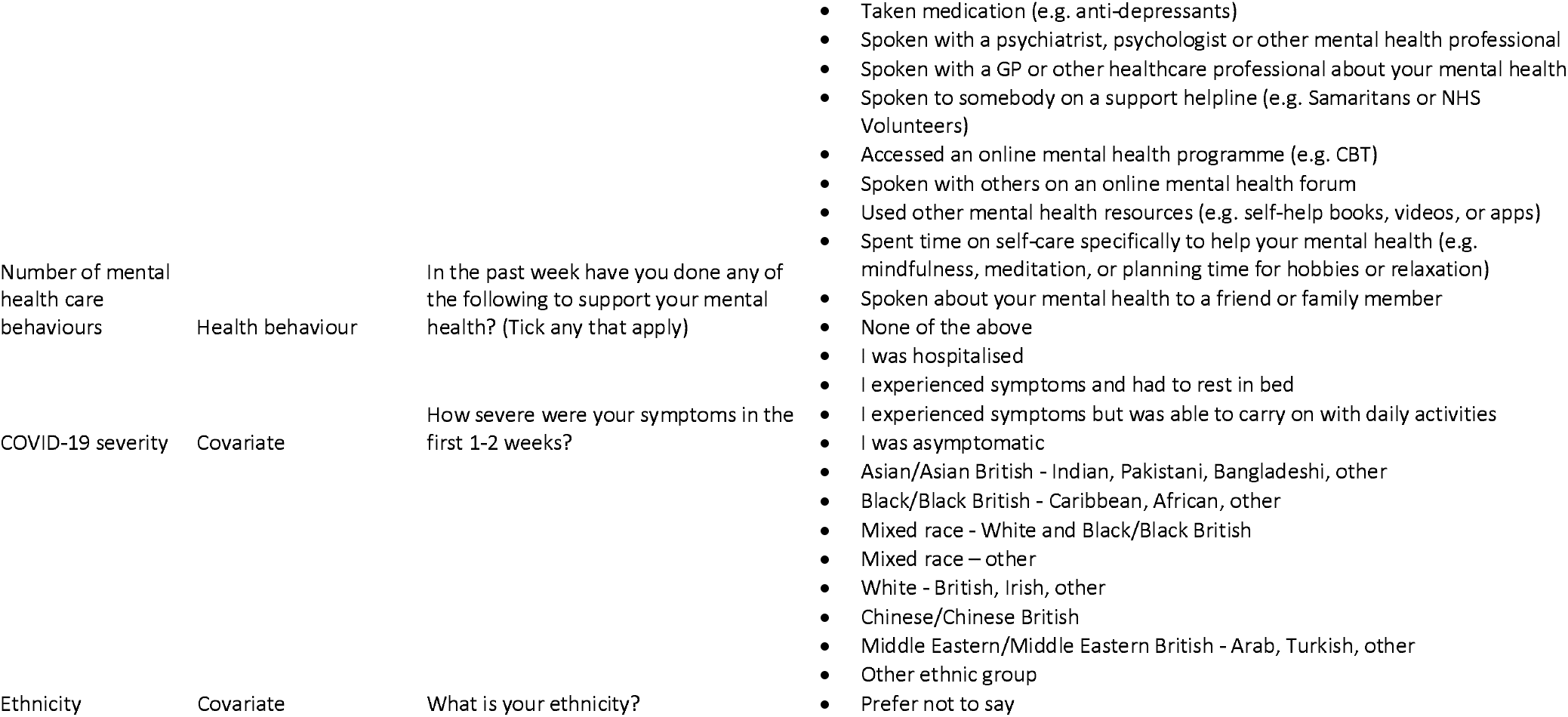
Wording of study developed items

**Table S8.** Number of weeks prior to COVID-19 infection in which health behaviours were measured (N = 1,811)

|  | Prop. |
| --- | --- |
| 2 | 26.06% |
| 3 | 24.02% |
| 4 | 24.68% |
| 5 | 21.76% |
| 6 | 3.48% |

**Table S9.** Weighted and unweighted sample characteristics (N = 1,811)

|  |  | Unweighted<br>Prop. | Weighted<br>Prop. |
| --- | --- | --- | --- |
| **Long COVID variables** |  |  |  |
| Long COVID status |  |  |  |
|  | Unsure | 12.70% | 12.04% |
|  | Yes, but not formally diagnosed | 12.31% | 16.68% |
|  | Yes, formally diagnosed | 3.87% | 3.80% |
| Presence of specific long COVID symptoms |  |  |  |
|  | Mobility (e.g., walking or climbing steps) | 43.02% | 44.92% |
|  | Cognition (e.g., remembering or concentrating) | 59.27% | 54.23% |
|  | Self-care (e.g., washing all over or dressing) | 10.71% | 12.10% |
| **Health behaviours in month prior to COVID-19 infection** |  |  |  |
| Exercise frequency (ref no exercise) |  |  |  |
|  | <30 min to 2 hours | 67.05% | 63.39% |
|  | 3+ hours | 25.30% | 27.72% |
| Days of fresh air (> 15 minutes) |  | 5.79 (1.91) | 5.63 (2.07) |
| Sleep quality (ref very good/good) |  |  |  |
|  | Average | 41.52% | 43.80% |
|  | Not good/very poor | 26.95% | 26.06% |
| Smoking (ref non-smoker) |  | 5.91% | 7.49% |
| More than 14 weekly alcoholic drinks (ref =< 14) |  | 9.94% | 10.56% |
| Number of mental health care behaviours |  | 0.74 (1.05) | 0.63 (1.01) |
| **COVID-19 infection variables** |  |  |  |
| COVID-19 infection severity in first two weeks (ref asymptomatic) |  |  |  |
|  | Mild | 38.27% | 41.11% |
|  | Moderate | 51.77% | 48.22% |
|  | Severe | 2.40% | 2.95% |
| Dominant strain in the UK at time of COVID-19 infection (ref original variant) |  |  |  |
|  | Alpha (1 November 2020 to 30 June 2021) | 41.58% | 38.36% |
|  | Delta (1 July to 30 November 2021) | 42.19% | 45.04% |
| **Socio-demographics** |  |  |  |
| Female (ref male) |  |  |  |
|  |  | 75.59% | 51.82% |
| Age (ref 60+) |  |  |  |
|  | 45-59 | 40.70% | 31.76% |
|  | 30-44 | 23.91% | 20.36% |
|  | 18-29 | 5.96% | 10.67% |
| Ethnic minority groups (ref White) |  | 3.64% | 6.12% |
| Education (ref degree or higher) |  |  |  |
|  | A-levels or vocational | 17.01% | 31.97% |
|  | Up to GCSE | 13.09% | 30.68% |
| Low household income (<£30,000) |  | 34.54% | 42.55% |
| Employed (ref not employed) |  | 67.59% | 59.42% |
| Key worker |  | 27.33% | 24.94% |
| Crowded household |  | 9.28% | 11.64% |
| Living arrangement (ref alone) |  |  |  |
|  | With others, not with children | 52.24% | 54.69% |
|  | With others, including children | 31.36% | 27.94% |
| Live in a rural area (ref urban) |  | 20.04% | 20.11% |
| **Pre-existing conditions** |  |  |  |
| Long-term physical health condition (ref none) |  |  |  |
|  |  | 38.87% | 43.97% |
| Long-term mental health condition (ref none) |  |  |  |
|  |  | 15.24% | 14.96% |

Note. Data in the weighted sample were weighted to the proportions of gender, age, ethnicity, country, and education obtained from the Office for National Statistics. GCSE refers to General Certificate of Secondary Education.

**Table S10.** Sensitivity analysis: logistic regressions predicting self-reported long COVID from health behaviours, with participants who were ‘unsure’ whether they had had long COVID in the case group (N = 1,811), weighted

|  |  | Self-reported long COVID |  |  |  |  |  |
| --- | --- | --- | --- | --- | --- | --- | --- |
|  |  | OR | SE | T | P | 95% CI | 95% CI |
| **Model 1** |  |  |  |  |  |  |  |
| Exercise frequency (ref no exercise) |  |  |  |  |  |  |  |
|  | <30 min to 2 hours | 0.75 | 0.22 | -0.97 | 0.33 | 0.42 | 1.35 |
|  | 3+ hours | 0.54 | 0.18 | -1.84 | 0.07 | 0.28 | 1.04 |
| Days of fresh air (> 15 minutes) |  |  |  |  |  |  |  |
|  |  | 0.98 | 0.04 | -0.42 | 0.67 | 0.90 | 1.07 |
| Sleep quality (ref very good/good) |  |  |  |  |  |  |  |
|  | Average | **1.86** | **0.35** | **3.31** | **<0.01** | **1.29** | **2.69** |
|  | Not good/very poor | **2.63** | **0.57** | **4.48** | **<0.01** | **1.72** | **4.01** |
| Smoking (ref non-smoker) |  |  |  |  |  |  |  |
|  |  | **2.08** | **0.74** | **2.07** | **0.04** | **1.04** | **4.17** |
| More than 14 weekly alcoholic drinks (ref =< 14) |  |  |  |  |  |  |  |
|  |  | 0.74 | 0.18 | -1.24 | 0.22 | 0.45 | 1.20 |
| Number of mental health care behaviours |  |  |  |  |  |  |  |
|  |  | 1.04 | 0.07 | 0.61 | 0.54 | 0.91 | 1.20 |
| **Model 2** |  |  |  |  |  |  |  |
| Exercise frequency (ref no exercise) |  |  |  |  |  |  |  |
|  | <30 min to 2 hours | 0.68 | 0.23 | -1.14 | 0.25 | 0.35 | 1.32 |
|  | 3+ hours | 0.49 | 0.18 | -1.95 | 0.05 | 0.24 | 1.00 |
| Days of fresh air (> 15 minutes) |  |  |  |  |  |  |  |
|  |  | 1.01 | 0.05 | 0.22 | 0.83 | 0.92 | 1.11 |
| Sleep quality (ref very good/good) |  |  |  |  |  |  |  |
|  | Average | **1.88** | **0.38** | **3.17** | **<0.01** | **1.27** | **2.78** |
|  | Not good/very poor | **2.44** | **0.53** | **4.09** | **<0.01** | **1.59** | **3.74** |
| Smoking (ref non-smoker) |  |  |  |  |  |  |  |
|  |  | **2.33** | **0.90** | **2.18** | **0.03** | **1.09** | **4.99** |
| More than 14 weekly alcoholic drinks (ref =< 14) |  |  |  |  |  |  |  |
|  |  | 0.84 | 0.22 | -0.67 | 0.50 | 0.49 | 1.41 |
| Number of mental health care behaviours |  |  |  |  |  |  |  |
|  |  | 0.98 | 0.07 | -0.29 | 0.77 | 0.85 | 1.12 |
| **Model 3** |  |  |  |  |  |  |  |
| Exercise frequency (ref no exercise) |  |  |  |  |  |  |  |
|  | <30 min to 2 hours | 0.72 | 0.23 | -1.02 | 0.31 | 0.38 | 1.36 |
|  | 3+ hours | **0.48** | **0.17** | **-2.05** | **0.04** | **0.24** | **0.97** |
| Days of fresh air (> 15 minutes) |  |  |  |  |  |  |  |
|  |  | 1.03 | 0.05 | 0.57 | 0.57 | 0.94 | 1.13 |
| Sleep quality (ref very good/good) |  |  |  |  |  |  |  |
|  | Average | **1.84** | **0.37** | **3.08** | **<0.01** | **1.25** | **2.72** |
|  | Not good/very poor | **2.28** | **0.50** | **3.75** | **<0.01** | **1.48** | **3.51** |
| Smoking (ref non-smoker) |  |  |  |  |  |  |  |
|  |  | **1.95** | **0.61** | **2.13** | **0.03** | **1.06** | **3.61** |
| More than 14 weekly alcoholic drinks (ref =< 14) |  |  |  |  |  |  |  |
|  |  | 0.83 | 0.23 | -0.67 | 0.51 | 0.48 | 1.44 |
| Number of mental health care behaviours |  |  |  |  |  |  |  |
|  |  | 1.02 | 0.07 | 0.33 | 0.74 | 0.89 | 1.18 |
| **Model 4** |  |  |  |  |  |  |  |
| Exercise frequency (ref no exercise) |  |  |  |  |  |  |  |
|  | <30 min to 2 hours | 0.69 | 0.23 | -1.11 | 0.27 | 0.36 | 1.33 |
|  | 3+ hours | **0.47** | **0.17** | **-2.09** | **0.04** | **0.23** | **0.95** |
| Days of fresh air (> 15 minutes) |  |  |  |  |  |  |  |
|  |  | 1.03 | 0.05 | 0.59 | 0.55 | 0.94 | 1.13 |
| Sleep quality (ref very good/good) |  |  |  |  |  |  |  |
|  | Average | **1.84** | **0.37** | **3.08** | **<0.01** | **1.25** | **2.72** |
|  | Not good/very poor | **2.29** | **0.51** | **3.77** | **<0.01** | **1.49** | **3.54** |
| Smoking (ref non-smoker) |  |  |  |  |  |  |  |
|  |  | **1.90** | **0.57** | **2.12** | **0.03** | **1.05** | **3.43** |
| More than 14 weekly alcoholic drinks (ref =< 14) |  |  |  |  |  |  |  |
|  |  | 0.82 | 0.23 | -0.71 | 0.48 | 0.48 | 1.41 |
| Number of mental health care behaviours |  |  |  |  |  |  |  |
|  |  | 1.04 | 0.08 | 0.56 | 0.58 | 0.90 | 1.21 |

Note. Data were weighted to the proportions of gender, age, ethnicity, country, and education obtained from the Office for National Statistics. Model 1 included only health behaviours (in the same model), Model 2 additionally adjusted for COVID-19 infection variables, Model 3 additionally adjusted for socio-demographic characteristics, and Model 4 additionally adjusted for pre-existing conditions.

**Table S11.** Sensitivity analysis: logistic regressions predicting the development of difficulty with mobility from health behaviours with participants who were ‘unsure’ whether they had had long COVID in the case group (N = 523), weighted

|  |  | Difficulty with mobility |  |  |  |  |  |
| --- | --- | --- | --- | --- | --- | --- | --- |
|  |  | OR | SE | T | P | 95% CI | 95% CI |
| **Model 1** |  |  |  |  |  |  |  |
| Exercise frequency (ref no exercise) |  |  |  |  |  |  |  |
|  | <30 min to 2 hours | 1.28 | 0.59 | 0.53 | 0.59 | 0.52 | 3.16 |
|  | 3+ hours | 0.99 | 0.51 | -0.02 | 0.99 | 0.36 | 2.71 |
| Days of fresh air (> 15 minutes) |  | 0.90 | 0.06 | -1.53 | 0.13 | 0.79 | 1.03 |
| Sleep quality (ref very good/good) |  |  |  |  |  |  |  |
|  | Average | 1.33 | 0.44 | 0.86 | 0.39 | 0.70 | 2.53 |
|  | Not good/very poor | 1.64 | 0.60 | 1.36 | 0.17 | 0.80 | 3.36 |
| Smoking (ref non-smoker) |  | 0.53 | 0.26 | -1.28 | 0.20 | 0.21 | 1.39 |
| More than 14 weekly alcoholic drinks (ref =< 14) |  | **2.33** | **1.00** | **1.96** | **0.05** | **1.00** | **5.42** |
| Number of mental health care behaviours |  | 1.18 | 0.15 | 1.36 | 0.17 | 0.93 | 1.50 |
| **Model 2** |  |  |  |  |  |  |  |
| Exercise frequency (ref no exercise) |  |  |  |  |  |  |  |
|  | <30 min to 2 hours | 1.23 | 0.58 | 0.44 | 0.66 | 0.49 | 3.11 |
|  | 3+ hours | 0.93 | 0.49 | -0.13 | 0.90 | 0.33 | 2.60 |
| Days of fresh air (> 15 minutes) |  | 0.92 | 0.06 | -1.21 | 0.23 | 0.80 | 1.05 |
| Sleep quality (ref very good/good) |  |  |  |  |  |  |  |
|  | Average | 1.35 | 0.48 | 0.85 | 0.40 | 0.68 | 2.70 |
|  | Not good/very poor | 1.63 | 0.63 | 1.26 | 0.21 | 0.76 | 3.48 |
| Smoking (ref non-smoker) |  | 0.57 | 0.28 | -1.14 | 0.25 | 0.21 | 1.51 |
| More than 14 weekly alcoholic drinks (ref =< 14) |  | **2.66** | **1.28** | **2.04** | **0.04** | **1.04** | **6.81** |
| Number of mental health care behaviours |  | 1.14 | 0.15 | 0.98 | 0.33 | 0.88 | 1.47 |
| **Model 3** |  |  |  |  |  |  |  |
| Exercise frequency (ref no exercise) |  |  |  |  |  |  |  |
|  | <30 min to 2 hours | 1.08 | 0.48 | 0.18 | 0.86 | 0.45 | 2.60 |
|  | 3+ hours | 0.68 | 0.35 | -0.76 | 0.45 | 0.25 | 1.85 |
| Days of fresh air (> 15 minutes) |  | 0.96 | 0.06 | -0.56 | 0.58 | 0.85 | 1.10 |
| Sleep quality (ref very good/good) |  |  |  |  |  |  |  |
|  | Average | 1.32 | 0.47 | 0.78 | 0.44 | 0.66 | 2.66 |
|  | Not good/very poor | 1.51 | 0.61 | 1.01 | 0.31 | 0.68 | 3.34 |
| Smoking (ref non-smoker) |  | 0.64 | 0.34 | -0.84 | 0.40 | 0.22 | 1.83 |
| More than 14 weekly alcoholic drinks (ref =< 14) |  | **2.80** | **1.45** | **1.99** | **0.05** | **1.01** | **7.72** |
| Number of mental health care behaviours |  | 1.29 | 0.18 | 1.87 | 0.06 | 0.99 | 1.69 |
| **Model 4** |  |  |  |  |  |  |  |
| Exercise frequency (ref no exercise) |  |  |  |  |  |  |  |
|  | <30 min to 2 hours | 0.96 | 0.44 | -0.09 | 0.93 | 0.39 | 2.37 |
|  | 3+ hours | 0.61 | 0.32 | -0.94 | 0.35 | 0.22 | 1.70 |
| Days of fresh air (> 15 minutes) |  | 0.97 | 0.07 | -0.48 | 0.63 | 0.85 | 1.10 |
| Sleep quality (ref very good/good) |  |  |  |  |  |  |  |
|  | Average | 1.26 | 0.45 | 0.65 | 0.52 | 0.63 | 2.54 |
|  | Not good/very poor | 1.52 | 0.62 | 1.01 | 0.31 | 0.68 | 3.40 |
| Smoking (ref non-smoker) |  | 0.59 | 0.33 | -0.95 | 0.34 | 0.20 | 1.77 |
| More than 14 weekly alcoholic drinks (ref =< 14) |  | 2.60 | 1.34 | 1.85 | 0.06 | 0.94 | 7.15 |
| Number of mental health care behaviours |  | **1.38** | **0.20** | **2.18** | **0.03** | **1.03** | **1.85** |

Note. Data were weighted to the proportions of gender, age, ethnicity, country, and education obtained from the Office for National Statistics. Model 1 included only health behaviours (in the same model), Model 2 additionally adjusted for COVID-19 infection variables, Model 3 additionally adjusted for socio-demographic characteristics, and Model 4 additionally adjusted for pre-existing conditions.

**Table S12.** Sensitivity analysis: logistic regressions predicting the development of difficulty with cognition from health behaviours, with participants who were ‘unsure’ whether they had had long COVID in the case group (N = 523), weighted

|  |  | Difficulty with cognition |  |  |  |  |  |
| --- | --- | --- | --- | --- | --- | --- | --- |
|  |  | OR | SE | T | P | 95% CI | 95% CI |
| **Model 1** |  |  |  |  |  |  |  |
| Exercise frequency (ref no exercise) |  |  |  |  |  |  |  |
|  | <30 min to 2 hours | 2.28 | 1.00 | 1.88 | 0.06 | 0.96 | 5.39 |
|  | 3+ hours | 2.40 | 1.19 | 1.77 | 0.08 | 0.91 | 6.32 |
| Days of fresh air (> 15 minutes) |  |  |  |  |  |  |  |
|  |  | 0.96 | 0.07 | -0.62 | 0.54 | 0.84 | 1.10 |
| Sleep quality (ref very good/good) |  |  |  |  |  |  |  |
|  | Average | 1.18 | 0.38 | 0.53 | 0.60 | 0.63 | 2.22 |
|  | Not good/very poor | **2.60** | **0.95** | **2.62** | **0.01** | **1.27** | **5.32** |
| Smoking (ref non-smoker) |  |  |  |  |  |  |  |
|  |  | 1.26 | 0.70 | 0.41 | 0.68 | 0.42 | 3.77 |
| More than 14 weekly alcoholic drinks (ref =< 14) |  |  |  |  |  |  |  |
|  |  | 1.11 | 0.49 | 0.24 | 0.81 | 0.47 | 2.65 |
| Number of mental health care behaviours |  |  |  |  |  |  |  |
|  |  | 1.27 | 0.25 | 1.23 | 0.22 | 0.87 | 1.86 |
| **Model 2** |  |  |  |  |  |  |  |
| Exercise frequency (ref no exercise) |  |  |  |  |  |  |  |
|  | <30 min to 2 hours | 1.88 | 0.83 | 1.43 | 0.15 | 0.79 | 4.45 |
|  | 3+ hours | 1.94 | 0.98 | 1.31 | 0.19 | 0.72 | 5.23 |
| Days of fresh air (> 15 minutes) |  |  |  |  |  |  |  |
|  |  | 0.97 | 0.07 | -0.45 | 0.66 | 0.85 | 1.11 |
| Sleep quality (ref very good/good) |  |  |  |  |  |  |  |
|  | Average | 1.07 | 0.35 | 0.21 | 0.84 | 0.57 | 2.03 |
|  | Not good/very poor | **2.28** | **0.86** | **2.20** | **0.03** | **1.09** | **4.77** |
| Smoking (ref non-smoker) |  |  |  |  |  |  |  |
|  |  | 1.39 | 0.89 | 0.51 | 0.61 | 0.40 | 4.86 |
| More than 14 weekly alcoholic drinks (ref =< 14) |  |  |  |  |  |  |  |
|  |  | 1.29 | 0.66 | 0.50 | 0.62 | 0.47 | 3.52 |
| Number of mental health care behaviours |  |  |  |  |  |  |  |
|  |  | 1.22 | 0.25 | 0.96 | 0.34 | 0.81 | 1.82 |
| **Model 3** |  |  |  |  |  |  |  |
| Exercise frequency (ref no exercise) |  |  |  |  |  |  |  |
|  | <30 min to 2 hours | 1.59 | 0.78 | 0.95 | 0.34 | 0.61 | 4.15 |
|  | 3+ hours | 1.47 | 0.79 | 0.72 | 0.47 | 0.51 | 4.20 |
| Days of fresh air (> 15 minutes) |  |  |  |  |  |  |  |
|  |  | 0.95 | 0.07 | -0.69 | 0.49 | 0.83 | 1.10 |
| Sleep quality (ref very good/good) |  |  |  |  |  |  |  |
|  | Average | 1.12 | 0.37 | 0.33 | 0.74 | 0.58 | 2.15 |
|  | Not good/very poor | 2.14 | 0.85 | 1.91 | 0.06 | 0.98 | 4.67 |
| Smoking (ref non-smoker) |  |  |  |  |  |  |  |
|  |  | 1.27 | 0.62 | 0.49 | 0.62 | 0.49 | 3.30 |
| More than 14 weekly alcoholic drinks (ref =< 14) |  |  |  |  |  |  |  |
|  |  | 1.46 | 0.72 | 0.76 | 0.45 | 0.55 | 3.83 |
| Number of mental health care behaviours |  |  |  |  |  |  |  |
|  |  | 1.38 | 0.29 | 1.51 | 0.13 | 0.91 | 2.09 |
| **Model 4** |  |  |  |  |  |  |  |
| Exercise frequency (ref no exercise) |  |  |  |  |  |  |  |
|  | <30 min to 2 hours | 1.61 | 0.80 | 0.97 | 0.33 | 0.61 | 4.26 |
|  | 3+ hours | 1.49 | 0.81 | 0.74 | 0.46 | 0.52 | 4.31 |
| Days of fresh air (> 15 minutes) |  |  |  |  |  |  |  |
|  |  | 0.96 | 0.07 | -0.62 | 0.53 | 0.83 | 1.10 |
| Sleep quality (ref very good/good) |  |  |  |  |  |  |  |
|  | Average | 1.11 | 0.37 | 0.31 | 0.76 | 0.57 | 2.15 |
|  | Not good/very poor | 2.14 | 0.84 | 1.94 | 0.05 | 0.99 | 4.61 |
| Smoking (ref non-smoker) |  |  |  |  |  |  |  |
|  |  | 1.24 | 0.61 | 0.43 | 0.66 | 0.47 | 3.24 |
| More than 14 weekly alcoholic drinks (ref =< 14) |  |  |  |  |  |  |  |
|  |  | 1.40 | 0.69 | 0.67 | 0.50 | 0.53 | 3.69 |
| Number of mental health care behaviours |  |  |  |  |  |  |  |
|  |  | 1.33 | 0.29 | 1.32 | 0.19 | 0.87 | 2.05 |

Note. Data were weighted to the proportions of gender, age, ethnicity, country, and education obtained from the Office for National Statistics. Model 1 included only health behaviours (in the same model), Model 2 additionally adjusted for COVID-19 infection variables, Model 3 additionally adjusted for socio-demographic characteristics, and Model 4 additionally adjusted for pre-existing conditions.

**Table S13.** Sensitivity analysis: logistic regressions predicting the development of difficulty with self-care from health behaviours, with participants who were ‘unsure’ whether they had had long COVID in the case group (N = 512), weighted

|  | Difficulty with self-care |  |  |  |  |  |
| --- | --- | --- | --- | --- | --- | --- |
|  | OR | SE | T | P | 95% CI | 95% CI |
| **Model 1** |  |  |  |  |  |  |
| Exercise frequency (ref no exercise) |  |  |  |  |  |  |
| <30 min to 2 hours | 0.88 | 0.51 | -0.22 | 0.83 | 0.28 | 2.73 |
| 3+ hours | 0.48 | 0.36 | -0.98 | 0.33 | 0.11 | 2.08 |
| Days of fresh air (> 15 minutes) | 0.85 | 0.07 | -1.93 | 0.05 | 0.72 | 1.00 |
| Sleep quality (ref very good/good) |  |  |  |  |  |  |
| Average | 1.94 | 1.11 | 1.16 | 0.24 | 0.63 | 5.95 |
| Not good/very poor | **3.46** | **1.97** | **2.18** | **0.03** | **1.13** | **10.56** |
| Smoking (ref non-smoker) | 1.40 | 0.73 | 0.64 | 0.52 | 0.50 | 3.87 |
| More than 14 weekly alcoholic drinks (ref =< 14) | **3.21** | **1.90** | **1.97** | **0.05** | **1.00** | **10.24** |
| Number of mental health care behaviours | 1.09 | 0.18 | 0.51 | 0.61 | 0.79 | 1.49 |
| **Model 2** |  |  |  |  |  |  |
| Exercise frequency (ref no exercise) |  |  |  |  |  |  |
| <30 min to 2 hours | 0.95 | 0.69 | -0.07 | 0.95 | 0.23 | 3.97 |
| 3+ hours | 0.49 | 0.43 | -0.80 | 0.42 | 0.09 | 2.75 |
| Days of fresh air (> 15 minutes) | 0.85 | 0.08 | -1.73 | 0.08 | 0.71 | 1.02 |
| Sleep quality (ref very good/good) |  |  |  |  |  |  |
| Average | 1.77 | 1.14 | 0.89 | 0.38 | 0.50 | 6.26 |
| Not good/very poor | 3.28 | 2.23 | 1.75 | 0.08 | 0.87 | 12.41 |
| Smoking (ref non-smoker) | 1.39 | 0.80 | 0.57 | 0.57 | 0.45 | 4.32 |
| More than 14 weekly alcoholic drinks (ref =< 14) | **3.52** | **2.25** | **1.97** | **0.05** | **1.01** | **12.31** |
| Number of mental health care behaviours | 1.01 | 0.17 | 0.08 | 0.94 | 0.73 | 1.41 |
| **Model 3** |  |  |  |  |  |  |
| Exercise frequency (ref no exercise) |  |  |  |  |  |  |
| <30 min to 2 hours | 0.75 | 0.51 | -0.43 | 0.67 | 0.20 | 2.81 |
| 3+ hours | 0.32 | 0.23 | -1.55 | 0.12 | 0.08 | 1.35 |
| Days of fresh air (> 15 minutes) | 0.89 | 0.08 | -1.24 | 0.22 | 0.74 | 1.07 |
| Sleep quality (ref very good/good) |  |  |  |  |  |  |
| Average | 1.62 | 1.24 | 0.64 | 0.52 | 0.37 | 7.21 |
| Not good/very poor | 2.24 | 1.81 | 1.00 | 0.32 | 0.46 | 10.90 |
| Smoking (ref non-smoker) | 2.45 | 1.39 | 1.58 | 0.11 | 0.81 | 7.45 |
| More than 14 weekly alcoholic drinks (ref =< 14) | **5.90** | **3.77** | **2.78** | **0.01** | **1.68** | **20.66** |
| Number of mental health care behaviours | 1.01 | 0.20 | 0.07 | 0.94 | 0.68 | 1.51 |
| **Model 4** |  |  |  |  |  |  |
| Exercise frequency (ref no exercise) |  |  |  |  |  |  |
| <30 min to 2 hours | 0.46 | 0.29 | -1.23 | 0.22 | 0.13 | 1.58 |
| 3+ hours | **0.18** | **0.13** | **-2.33** | **0.02** | **0.04** | **0.76** |
| Days of fresh air (> 15 minutes) | 0.88 | 0.09 | -1.29 | 0.20 | 0.72 | 1.07 |
| Sleep quality (ref very good/good) |  |  |  |  |  |  |
| Average | 1.39 | 1.13 | 0.41 | 0.68 | 0.28 | 6.82 |
| Not good/very poor | 2.65 | 2.25 | 1.14 | 0.25 | 0.50 | 14.05 |
| Smoking (ref non-smoker) | 2.84 | 1.76 | 1.68 | 0.09 | 0.84 | 9.59 |
| More than 14 weekly alcoholic drinks (ref =< 14) | **5.24** | **3.52** | **2.46** | **0.01** | **1.40** | **19.58** |
| Number of mental health care behaviours | 1.12 | 0.26 | 0.47 | 0.64 | 0.71 | 1.75 |

Note. Data were weighted to the proportions of gender, age, ethnicity, country, and education obtained from the Office for National Statistics. Model 1 included only health behaviours (in the same model), Model 2 additionally adjusted for COVID-19 infection variables, Model 3 additionally adjusted for socio-demographic characteristics, and Model 4 additionally adjusted for pre-existing conditions.

**Table S14.**
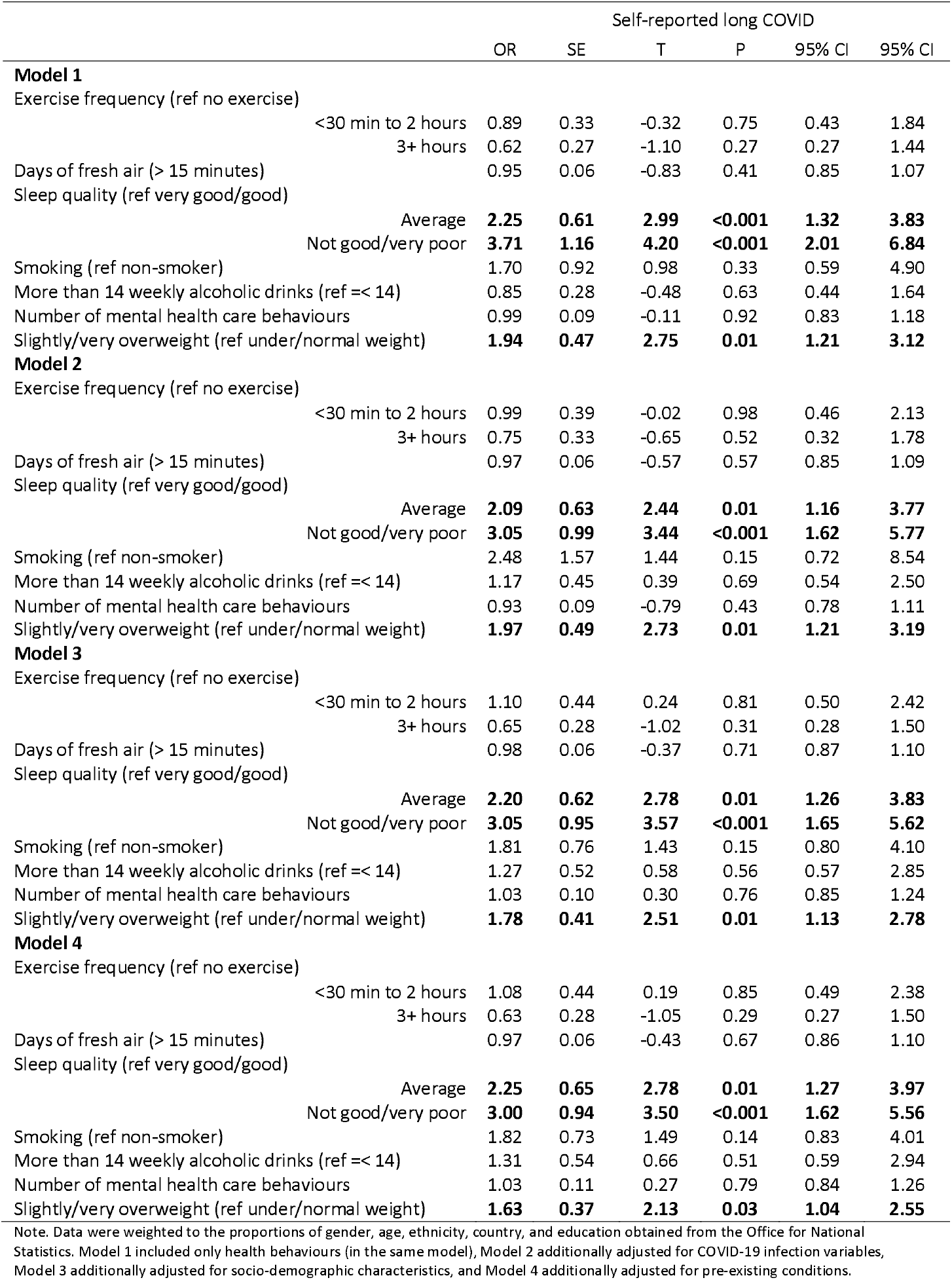
Sensitivity analysis: logistic regressions predicting the development of long COVID from health behaviours, including overweight/obesity status (N = 1,283) weighted

**Table S15.** Sensitivity analysis: logistic regressions predicting the development of difficulty with mobility from health behaviours, including overweight/obesity status (N = 234), weighted

|  | Difficulty with mobility |  |  |  |  |  |
| --- | --- | --- | --- | --- | --- | --- |
|  | OR | SE | T | P | 95% CI | 95% CI |
| **Model 1** |  |  |  |  |  |  |
| Exercise frequency (ref no exercise) |  |  |  |  |  |  |
| <30 min to 2 hours | 0.75 | 0.49 | -0.44 | 0.66 | 0.21 | 2.72 |
| 3+ hours | 0.64 | 0.48 | -0.59 | 0.55 | 0.15 | 2.78 |
| Days of fresh air (> 15 minutes) | 0.88 | 0.08 | -1.43 | 0.15 | 0.74 | 1.05 |
| Sleep quality (ref very good/good) |  |  |  |  |  |  |
| Average | 0.66 | 0.35 | -0.78 | 0.44 | 0.23 | 1.87 |
| Not good/very poor | 0.99 | 0.55 | -0.02 | 0.98 | 0.33 | 2.93 |
| Smoking (ref non-smoker) | 0.51 | 0.38 | -0.91 | 0.37 | 0.12 | 2.19 |
| More than 14 weekly alcoholic drinks (ref =< 14) | 1.78 | 1.10 | 0.93 | 0.35 | 0.53 | 5.98 |
| Number of mental health care behaviours | 1.09 | 0.18 | 0.51 | 0.61 | 0.78 | 1.51 |
| Slightly/very overweight (ref under/normal weight) | 1.40 | 0.63 | 0.75 | 0.46 | 0.58 | 3.38 |
| **Model 2** |  |  |  |  |  |  |
| Exercise frequency (ref no exercise) |  |  |  |  |  |  |
| <30 min to 2 hours | 0.69 | 0.48 | -0.53 | 0.59 | 0.18 | 2.71 |
| 3+ hours | 0.67 | 0.54 | -0.50 | 0.61 | 0.14 | 3.21 |
| Days of fresh air (> 15 minutes) | 0.90 | 0.08 | -1.12 | 0.26 | 0.76 | 1.08 |
| Sleep quality (ref very good/good) |  |  |  |  |  |  |
| Average | 0.63 | 0.31 | -0.92 | 0.36 | 0.24 | 1.68 |
| Not good/very poor | 0.84 | 0.46 | -0.32 | 0.75 | 0.29 | 2.47 |
| Smoking (ref non-smoker) | 0.74 | 0.52 | -0.44 | 0.66 | 0.19 | 2.92 |
| More than 14 weekly alcoholic drinks (ref =< 14) | 2.52 | 1.88 | 1.24 | 0.22 | 0.58 | 10.91 |
| Number of mental health care behaviours | 1.06 | 0.18 | 0.31 | 0.75 | 0.75 | 1.49 |
| Slightly/very overweight (ref under/normal weight) | 1.27 | 0.57 | 0.54 | 0.59 | 0.53 | 3.05 |
| **Model 3** |  |  |  |  |  |  |
| Exercise frequency (ref no exercise) |  |  |  |  |  |  |
| <30 min to 2 hours | 1.32 | 1.07 | 0.34 | 0.73 | 0.27 | 6.50 |
| 3+ hours | 0.74 | 0.69 | -0.32 | 0.75 | 0.12 | 4.61 |
| Days of fresh air (> 15 minutes) | 0.90 | 0.09 | -1.02 | 0.31 | 0.73 | 1.10 |
| Sleep quality (ref very good/good) |  |  |  |  |  |  |
| Average | 0.77 | 0.47 | -0.43 | 0.66 | 0.23 | 2.56 |
| Not good/very poor | 0.65 | 0.41 | -0.69 | 0.49 | 0.19 | 2.23 |
| Smoking (ref non-smoker) | 0.88 | 0.62 | -0.18 | 0.86 | 0.22 | 3.49 |
| More than 14 weekly alcoholic drinks (ref =< 14) | 4.45 | 3.46 | 1.92 | 0.06 | 0.97 | 20.45 |
| Number of mental health care behaviours | 1.29 | 0.24 | 1.36 | 0.17 | 0.89 | 1.87 |
| Slightly/very overweight (ref under/normal weight) | 1.46 | 0.68 | 0.80 | 0.42 | 0.58 | 3.66 |
| **Model 4** |  |  |  |  |  |  |
| Exercise frequency (ref no exercise) |  |  |  |  |  |  |
| <30 min to 2 hours | 1.33 | 1.03 | 0.37 | 0.71 | 0.29 | 6.10 |
| 3+ hours | 0.73 | 0.68 | -0.34 | 0.74 | 0.12 | 4.51 |
| Days of fresh air (> 15 minutes) | 0.90 | 0.10 | -1.01 | 0.31 | 0.73 | 1.11 |
| Sleep quality (ref very good/good) |  |  |  |  |  |  |
| Average | 0.73 | 0.46 | -0.50 | 0.62 | 0.21 | 2.51 |
| Not good/very poor | 0.63 | 0.41 | -0.71 | 0.48 | 0.18 | 2.25 |
| Smoking (ref non-smoker) | 0.88 | 0.63 | -0.17 | 0.86 | 0.22 | 3.58 |
| More than 14 weekly alcoholic drinks (ref =< 14) | 4.49 | 3.66 | 1.84 | 0.07 | 0.91 | 22.18 |
| Number of mental health care behaviours | 1.34 | 0.27 | 1.45 | 0.15 | 0.90 | 2.01 |
| Slightly/very overweight (ref under/normal weight) | 1.45 | 0.67 | 0.81 | 0.42 | 0.59 | 3.59 |

Note. Data were weighted to the proportions of gender, age, ethnicity, country, and education obtained from the Office for National Statistics. Model 1 included only health behaviours (in the same model), Model 2 additionally adjusted for COVID-19 infection variables, Model 3 additionally adjusted for socio-demographic characteristics, and Model 4 additionally adjusted for pre-existing conditions.

**Table S16.**
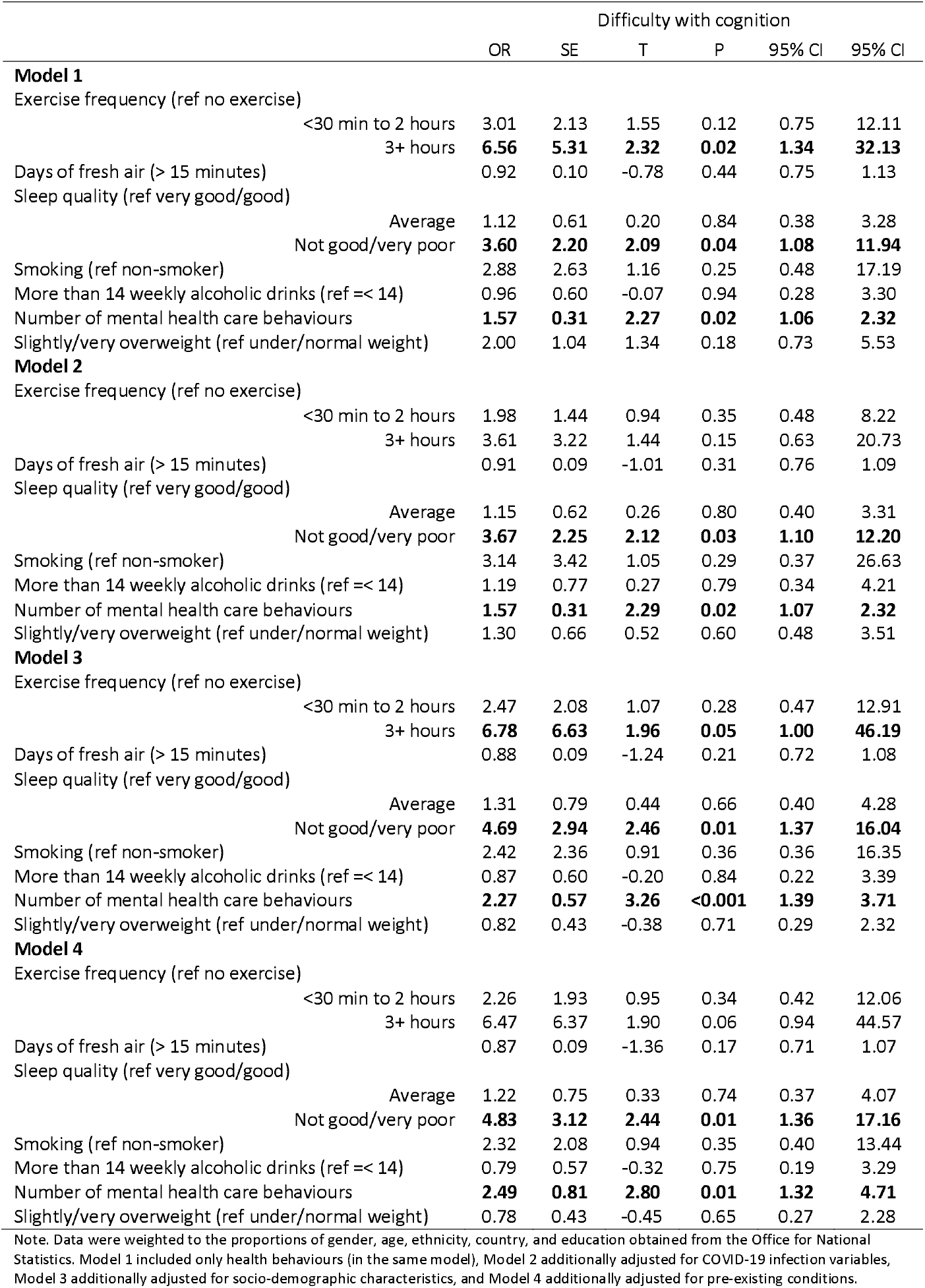
Sensitivity analysis: logistic regressions predicting the development of difficulty with cognition from health behaviours, including overweight/obesity status (N = 234) weighted

**Table S17.**
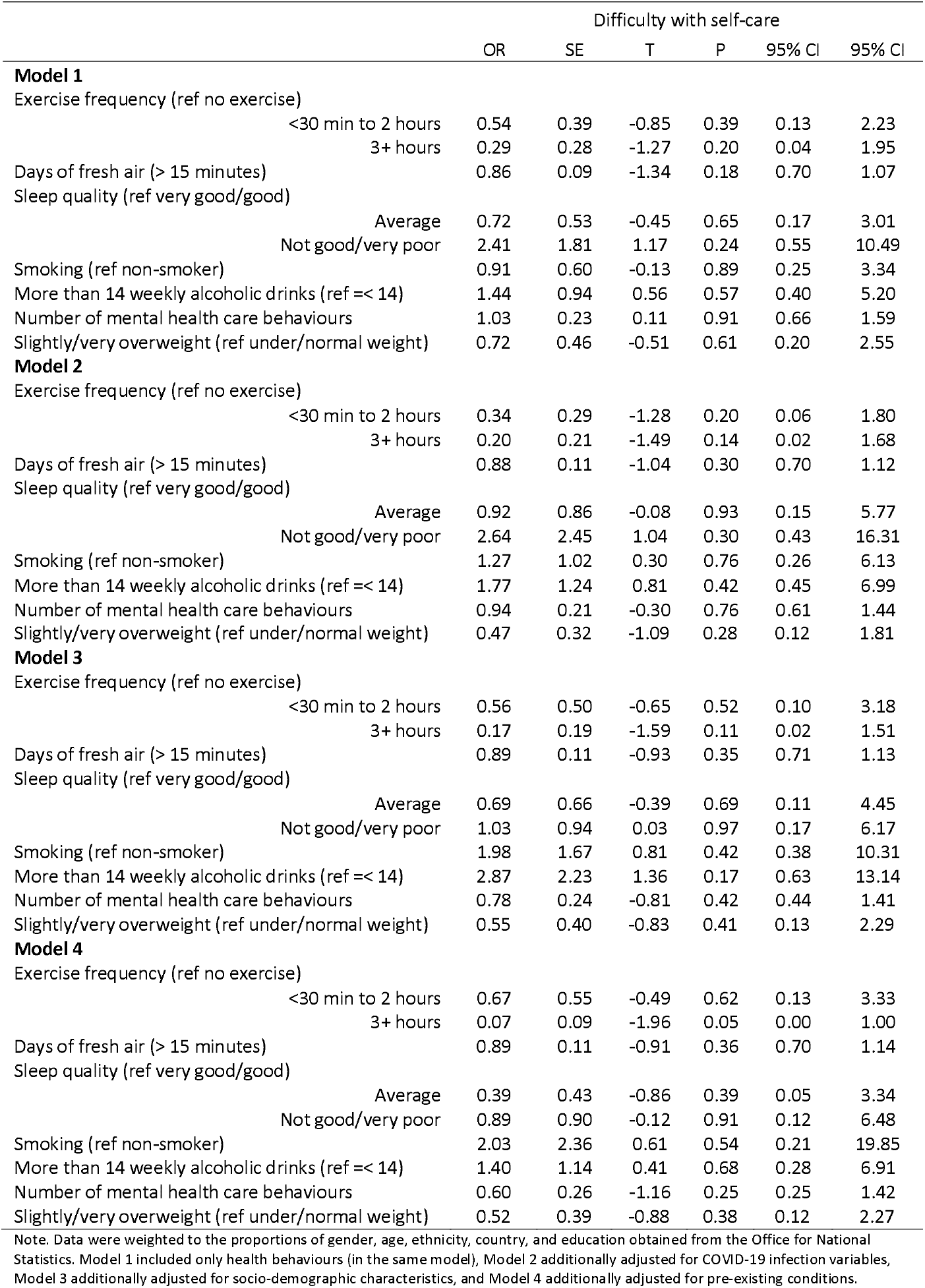
Sensitivity analysis: logistic regressions predicting the development of difficulty with self-care from health behaviours, including overweight/obesity status (N = 225) weighted

